# Excitation/inhibition imbalance and conversion to psychosis in the clinical high risk syndrome: Biophysical modeling finds reduced pyramidal cell excitability across EEG paradigms

**DOI:** 10.1101/2025.09.16.25335778

**Authors:** Julia Rodriguez-Sanchez, Daniel J. Hauke, Dimitris Pinotsis, Lioba C. S. Berndt, Hope Oloye, Spero C. Nicholas, Holly K. Hamilton, Brian J. Roach, Peter M. Bachman, Aysenil Belger, Ricardo E. Carrión, Erica Duncan, Jason K. Johannesen, Gregory A. Light, Margaret A. Niznikiewicz, Karl J. Friston, Jean Addington, Carrie E. Bearden, Kristin S. Cadenhead, Diana O. Perkins, Elaine F. Walker, Scott W. Woods, Tyrone D. Cannon, Rick A. Adams, Daniel H. Mathalon

**Affiliations:** Hawkes Institute, University College London, UK; Department of Psychology & Neuroscience, City St George’s, University of London, UK; Department of Psychology, Faculty of Health and Life Sciences, University of Exeter, UK; Institute of Cognitive Neuroscience, University College London, UK; Mental Health Service, San Francisco Veterans Affairs Health Care System, San Francisco, CA, USA; Northern California Institute for Research and Education, San Francisco, CA, USA; Mental Health Service, Minneapolis Veterans Affairs Health Care System, Minneapolis, MN, USA; Department of Psychiatry & Behavioral Sciences, University of Minnesota, Minneapolis, MN, USA; Department of Psychiatry & Behavioral Sciences, University of California, San Francisco, CA, USA; Department of Psychiatry, Boston Children’s Hospital, Boston, MA, USA; Department of Psychiatry, University of North Carolina at Chapel Hill, Chapel Hill, NC, USA; Division of Psychiatry Research, The Zucker Hillside Hospital, Northwell Health, Glen Oaks, NY, USA; Institute of Behavioral Science, Feinstein Institutes for Medical Research, North Shore-Long Island Jewish Health System, Manhasset, NY, USA; Mental Health Service, Atlanta Veterans Affairs Health Care System, Decatur, GA, USA; Department of Psychiatry and Behavioral Sciences, Emory University School of Medicine, Atlanta, GA, USA; Department of Psychiatry, Yale University, School of Medicine, New Haven, CT, USA; Department of Psychiatry, University of California, San Diego, La Jolla, CA, USA; Mental Health Service, Veterans Affairs San Diego Healthcare System, La Jolla, CA, USA; Department of Psychiatry, Harvard Medical School at Beth Israel Deaconess Medical Center and Massachusetts General Hospital, Boston, MA, USA; Mental Health Service, Veterans Affairs Boston Healthcare System, Brockton, MA, USA; Wellcome Trust Centre for Neuroimaging, Institute of Neurology, University College London, UK; Hotchkiss Brain Institute Department of Psychiatry, University of Calgary, AB, Canada; Department of Psychiatry and Biobehavioral Sciences, Semel Institute for Neuroscience and Human Behavior, University of California, Los Angeles, CA, USA; Department of Psychology, University of California, Los Angeles, CA, USA; Department of Psychology, Emory University, Atlanta, GA, USA; Department of Psychology, Yale University, New Haven, CT, USA

**Author notes:** Corresponding author: Julia Rodriguez-Sanchez 90 High Holborn London WC1V 6LJ United Kingdom. Drs. Rodriguez-Sanchez and Hauke are joint first authors. Drs. Adams and Mathalon are joint last authors.

**Keywords:** Clinical high risk for psychosis, dynamic causal modeling, biophysical modeling, MMN, P300, E/I balance

## Abstract

**Background:** Reduced mismatch negativity (MMN) and P300 event-related potential (ERP) components are widely replicated in schizophrenia and are also observed in individuals at clinical high risk for psychosis (CHR-P) who subsequently convert to psychosis. It is unknown whether they reflect changes in excitatory and/or inhibitory synaptic function, both implicated in schizophrenia and considered potential drug targets.

**Methods:** We analyzed baseline MMN and P300 ERPs from the NAPLS2 study, asking whether altered synaptic excitation, inhibition, or both could explain amplitude reductions in CHR-P (n=583). CHR-P participants who converted to psychosis (CHR-Converters; n=77) or remitted by 24-month follow-up (CHR-Remitters; n=94) were compared on MMN evoked by pitch+duration double-deviant tones and P300 elicited by target tones from passive and active auditory oddball paradigms, respectively. Biophysical modeling was used to infer (excitatory) pyramidal cell and (inhibitory) interneuron function from both MMN and P300 ERPs.

**Results:** MMN and P300 amplitude reductions in future CHR-Converters relative to CHR-Remitters were best explained by reduced pyramidal cell excitability (posterior probability P>.95 of a group-by-condition interaction effect). In simulations, reduced pyramidal cell excitability suppressed deviant and target ERPs. Within CHR-Converters, but not CHR-Remitters, more severe positive symptoms were associated with disinhibition of pyramidal cells (P>.99).

**Conclusions:** Results mirror previous findings in schizophrenia and suggest that reduced pyramidal cell excitability is present at baseline in future CHR-Converters, supporting the hypothesis that hypofunction of pyramidal cells is a primary pathology in schizophrenia, rather than a consequence of chronic illness. Positive symptoms among CHR-Converters may reflect compensatory downregulation of inhibition.

## Introduction

The onset of psychotic disorders like schizophrenia is commonly preceded by a prodrome marked by attenuated psychotic symptoms – such as perceptual abnormalities, unusual thought content, and disorganization of thought – which are of sub-threshold severity, duration, and/or conviction (1,2). Individuals meeting criteria for this “attenuated psychosis syndrome” are considered at clinical high risk for psychosis (CHR-P) (3). Meta-analytic estimates suggest that approximately 26% of CHR-P individuals develop a psychotic disorder within 2-3 years (4), while one of the largest scale studies conducted to date reported a smaller 2-year conversion rate of 16% (5). About 46% remit from the CHR-P syndrome over the same period (6). Crucially, no preventive treatments for CHR-P are available at present – a limitation attributed to the lack of mechanistic models of psychosis onset in CHR-P (7).

Two electroencephalography (EEG)-based measures have emerged as potential biomarkers of psychotic disorders: the mismatch negativity (MMN) and the P300 components of auditory event-related potentials (ERPs). They reflect neural responses to infrequent deviant stimuli relative to frequent standard stimuli. Specifically, the MMN is evoked in response to deviant sounds in the absence of attention, with participants typically instructed to ignore the sounds and focus on an incidental visual task (8). The MMN is thought to reflect auditory echoic memory and pre-attentive prediction error signaling (8–13). In contrast, the P300, which can be divided into P3a and P3b subcomponents, reflects attention-dependent responses to infrequent stimuli (9). P3a is elicited by ‘novel’ task-irrelevant distractor stimuli and reflects exogenous bottom-up attention switching, while P3b is elicited by infrequent task-relevant ‘target’ stimuli requiring a response and is related to endogenous top-down attentional allocation (14,15).

Reduced MNN (effect size [*ES*]=0.95-0.99) (10,16,17) and P3b (*ES*=0.74-0.85) (18,19) amplitudes are among the most robust findings in schizophrenia, and they have also been reported in CHR-P individuals (20,21). Moreover, reduced MMN (22–26) and target P300 (P3b) (27–30) have been associated with clinical outcomes in CHR-P. In the largest longitudinal study of CHR-P conducted to date (764 CHR-P, 280 healthy controls [HC]), Hamilton et al. (24,27) found reduced MMN and P3b amplitudes in CHR-P participants who converted to psychosis within 2 years (CHR-Converters). Specifically, MMN reductions were associated with future conversion to psychosis (24), distinguishing future CHR-Converters from non-converters (including both CHR-P with persistent symptoms [CHR-Persistent] and CHR-P who remitted from the CHR-P syndrome [CHR-Remitters]) and HC. P3b responses were decreased in CHR-Converters or CHR-Persistent, relative to CHR-Remitters, whose P3b responses were indistinguishable from HC (27).

An imbalance between excitatory and inhibitory (E/I) circuit function may be a fundamental pathophysiology in psychosis (31,32), and may also underlie MMN and P300 reductions in psychosis spectrum disorders (33,34). Hypofunction of NMDA receptors, induced via NMDA receptor antagonist challenge, reduces MMN amplitudes in non-human primate (35) and human (36–39) studies, and P300 amplitudes in human studies (36,37,40–42). In people with schizophrenia, MMN amplitude reductions are associated with lower brain glutamate and GABA levels assessed using proton-magnetic resonance spectroscopy (43), potentially implicating both reduced excitation and reduced inhibition. Nevertheless, the most direct evidence for disruptions in both excitatory and inhibitory function currently derives from post-mortem (44–46) and genetic (47) studies, as measuring excitation and inhibition in humans *in vivo* remains challenging (48,49).

Biophysical (i.e., neural mass) models and dynamic causal modeling (DCM) can, in principle, be used to infer excitatory and inhibitory cell function at the individual level from non-invasive EEG data (50), making it possible to study the synaptic pathology underlying MMN and P300 reductions. This is achieved by specifying how the activity of neuronal populations maps to the measured EEG signal and then inverting this mapping to estimate model parameters from the recorded data. Previous DCM studies of MMN and P300 found reduced excitatory (pyramidal) cell excitability in the deviant condition in people with schizophrenia (33,34). More recently, a DCM study of >100 people with schizophrenia reported reduced pyramidal cell excitability across four different EEG and functional magnetic resonance imaging (fMRI) paradigms (51). Interestingly, disinhibition (i.e., reduced interneuron function) correlated with abnormal auditory percepts. These findings may indicate a primary hypofunction of excitatory cells, with a compensatory downregulation of inhibition – to restore E/I balance – that then leads to positive symptoms (31,32,44). It is unknown, however, whether these changes are also seen in CHR-P or are unique to the chronic stage of psychosis due to progressive synaptic loss (52). If this mechanistic account of psychosis is true, pyramidal cell hypoexcitability should be present in CHR-P individuals, at least in those at greatest risk for conversion to psychosis.

Here, we apply a new model specifically designed for estimating excitatory and inhibitory cell function (53) to a large CHR-P dataset from the North American Prodrome Longitudinal Study 2 (NAPLS2) (54). We compare CHR-Converters to CHR-Remitters, a distinction crucial for identifying mechanisms of resilience and transition to psychosis. Indeed, while comparisons with HC are useful for characterizing alterations associated with the CHR-P state, the imperative is to find predictors that distinguish clinical outcomes among CHR-P individuals; especially given low conversion rates (4,5). We model two biomarkers that distinguish CHR-Converters and CHR-Remitters across passive (MMN) and active (target P300) auditory oddball paradigms (24,27) to probe the synaptic mechanisms underlying these differences. CHR-P participants who neither converted nor remitted exhibited a mixed biomarker profile – with MMN responses resembling CHR-Remitters and P3b responses more like CHR-Converters (24,27) – were excluded, ensuring consistent group comparisons in this cross-paradigm analysis. Drawing on previous findings in chronic schizophrenia (51), we hypothesized that CHR-Converters would exhibit reduced pyramidal cell excitability – reflecting early excitatory dysfunction as a key driver of illness onset – but a positive relationship between disinhibition and positive symptoms.

## Methods

### Participants

Participants were recruited from 8 university-based outpatient programs as part of the multisite, case-control NAPLS2 study (54,55). A total of n=764 individuals meeting criteria for the CHR-P syndrome based on the Structured Interview for Psychosis-Risk Syndromes (1) were followed for 24 months or until they converted to psychosis, with clinical assessments every 6 months. Symptoms were assessed using the Scale of Psychosis-Risk Symptoms, with CHR-Converters meeting Presence of Psychotic Symptoms criteria. Participants who completed the study but no longer met CHR-P criteria at the 24-month assessment were classified as CHR-Remitters. Participants who showed persistent symptoms without meeting conversion criteria at the 24-month assessment (CHR-Persistent) were excluded from this report, which focused on differences between the more ‘extreme’ outcomes: conversion and remission. Additional analyses including CHR-Persistent participants, and comparisons between CHR-Converters and age and sex matched HC, are provided in the **Supplement.**

Of the n=764 CHR-P, n=583 provided usable MMN and/or P300 data (see (24,27) for enrollment details and inclusion criteria). Among these, n=266 CHR-P non-converter participants did not complete the full 24-month follow-up and could therefore not be assigned to outcome groups with confidence. The final sample for analysis included n=77 CHR-Converters and n=94 CHR-Remitters for the MMN paradigm, and n=73 CHR-Converters and n=89 CHR-Remitters for the P300 paradigm (for demographics/clinical characteristics, see **Table 1**). At baseline, 23 CHR-Converters (29.9%) and 16 CHR-Remitters (17.9%) were receiving antipsychotic medication. Participants taking antipsychotics were included if their clinical history indicated that symptoms were within the CHR-P range (but not psychotic) at the time medication was started (55). To account for medication effects, group-level analyses were repeated 1) excluding medicated individuals and 2) including antipsychotic use at baseline as a covariate (**Supplement**).

**Table 1.**
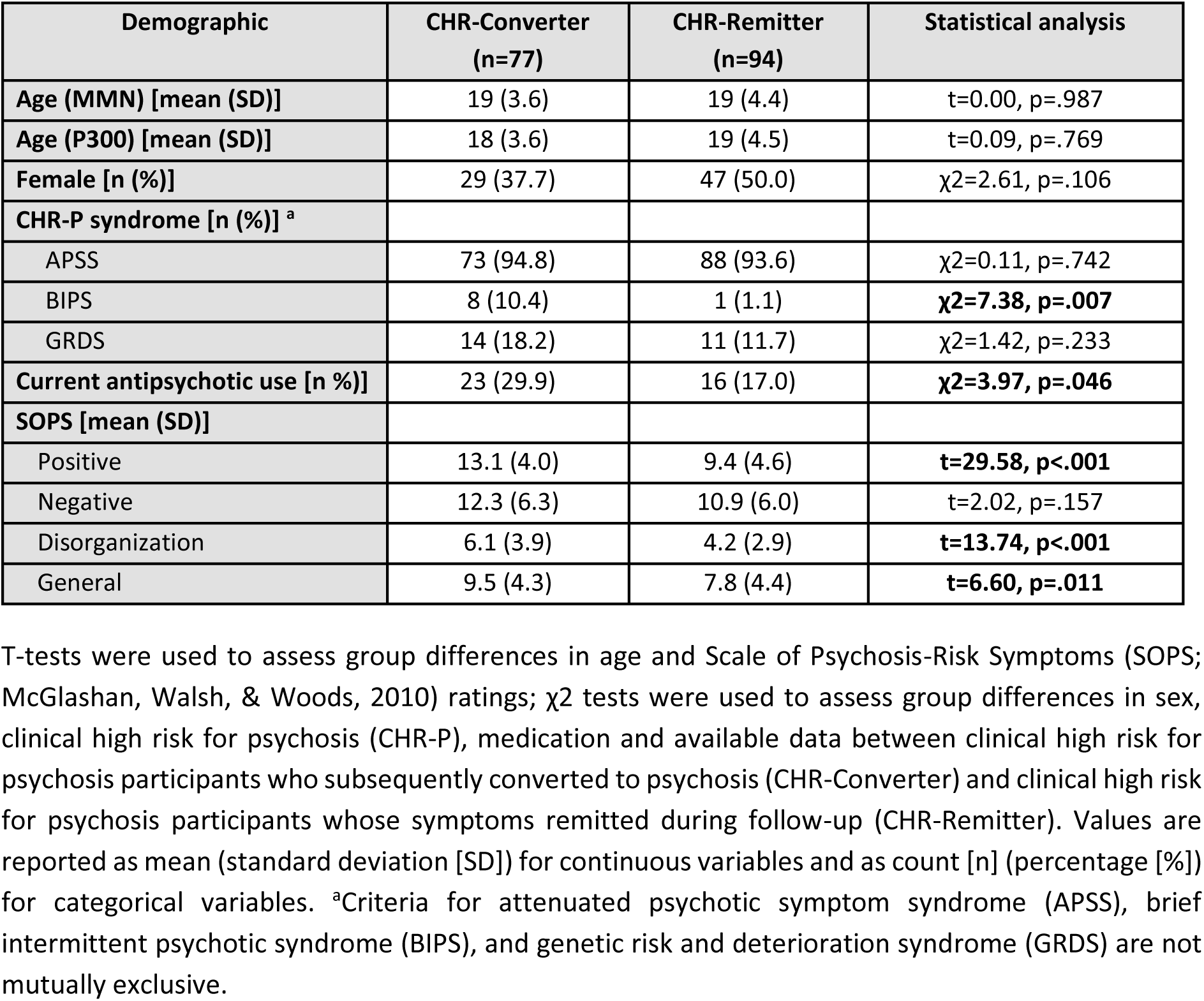
Demographic and clinical characteristics.

### Stimuli and task

Full descriptions of the experimental paradigms are reported elsewhere (24,27). Briefly, the MMN paradigm consisted of infrequent deviant tones presented in a pseudorandom sequence among frequent standard tones (633 Hz, 50 ms duration, 85%). The deviant tones included: a duration-deviant (633 Hz, 100 ms, 5%), a pitch-deviant (1000 Hz, 50 ms, 5%), and a duration+pitch double-deviant (1000 Hz, 100 ms, 5%). Participants were asked to ignore the auditory stimuli and engage in a visual oddball task. Only the MMN evoked by double-deviant tones was analyzed in this report, as it was the best predictor of conversion to psychosis, relative to the other deviant types (24).

The P300 paradigm consisted of a 3-stimulus auditory oddball task, with frequent standard tones (500 Hz, 80%) and randomly interspersed infrequent target tones (1,000 Hz, 10%) and task-irrelevant novel sounds (10%). Participants were instructed to respond only to the target tones by pressing a button with their preferred hand. Response accuracy was high (>98%) after excluding 4 CHR-P participants (1 CHR-Converter) who performed below chance levels (27). Only the P300 evoked by target stimuli (P3b) was analyzed in the current report, as it distinguished conversion from remission outcomes, whereas the P300 to novel stimuli (P3a) showed similar reduction in CHR-P relative to HC participants irrespective of clinical outcome (27).

### Data acquisition and preprocessing

EEG data were collected using a BioSemi ActiveTwo high-impedance recording system with either a 32-channel or a 64-channel electrode cap (4 study sites each) and digitized at 1024 Hz. Preprocessing is summarized in the **Supplement** and has been detailed previously (24,27). Averaged ERP waveforms were exported into SPM and downsampled to 250 Hz for sensor-level analyses. For modeling, data were further downsampled to 125 Hz to match the empirical residual autocorrelation with the residual autocorrelation assumed by the model, a procedure shown to improve model recovery (56).

### Sensor-level analysis

This study investigated the neurobiological mechanisms underlying MMN and P3b deficits, rather than the presence of previously established group effects (24,27). Prior sensor-level analyses of this dataset found significantly reduced P3b and MMN amplitudes in CHR-Converters compared to CHR-Remitters (24,27), although the MMN effect did not survive correction for multiple comparisons. For ease of reading, we illustrate sensor-level effects and the time window over which they were expressed using t-tests per timepoint (uncorrected) and report group differences assessed with independent sample t-tests (α=0.05, two-tailed, uncorrected). MMN was quantified at 6 frontocentral electrodes (F3, Fz, F4, C3, Cz, C4) by subtracting the standard from the double-deviant ERP and computing the mean amplitude 90-170 ms post-stimulus (24). P300 was quantified at electrode Pz by subtracting the standard from the target ERP and computing the mean amplitude 235-400 ms post-stimulus (14,27). Software details are reported in the **Supplement.**

### Network specification

Sources of the MMN during the passive oddball task have been well characterized (10,57–59) and include the primary auditory cortex, secondary auditory cortex (superior temporal gyrus, STG) and inferior frontal gyrus. A key difference in the active compared to the passive oddball paradigm is that participants must maintain top-down attention towards the target tones and respond via button presses (15). We therefore modeled the active (target P300) paradigm with a 6-region network comprising bilateral STG as auditory input regions, and regions of the dorsal attention network involved in top-down attention processing including the inferior frontal junction and inferior parietal sulcus ((60,61); see (53) and **Supplement**).

### Excitation/inhibition model

We modeled both paradigms using a new canonical microcircuit (CMC) model designed to estimate excitatory and inhibitory function (53). This neural mass model comprises two components: a latent state model characterizing the evolution of hidden neuronal states (voltage and current) over time, and a forward model mapping neuronal activity to the measured EEG signal. The state model is based on the microcircuit described in Bastos et al. (62), where each cortical source consists of interacting populations of superficial and deep pyramidal cells, spiny stellate cells and inhibitory interneurons, interconnected according to anatomical constraints (51,63). The forward model was based on equivalent current dipoles implemented in SPM.

Importantly, Hauke et al. (53) introduced two intrinsic connectivity parameters expressing the difference in E/I between experimental conditions: *B*^*gee*^ and *B*^*gii*^ (**Figure 1**). Here, the first and second subscripts indicate the origin and target of the intrinsic connections, respectively: ‘e’ for excitatory superficial pyramidal cells or ‘i’ for inhibitory interneurons. These parameters determine the synaptic gain or ‘excitability’ of excitatory and inhibitory cells, modeled implicitly through inhibitory self-connections. Since these connections are inhibitory, decreased *B*^*gee*^ or *B*^*gii*^ correspond to greater excitatory or inhibitory neuron excitability, respectively. Notably, we assumed that any group differences were not due to condition-specific changes in extrinsic (i.e., between-source) connections (which were fixed to the grand mean posterior), but instead reflected changes in the condition-specific rebalancing of E/I implicit in intrinsic (i.e., within-source) synaptic connections.

**Figure 1.**
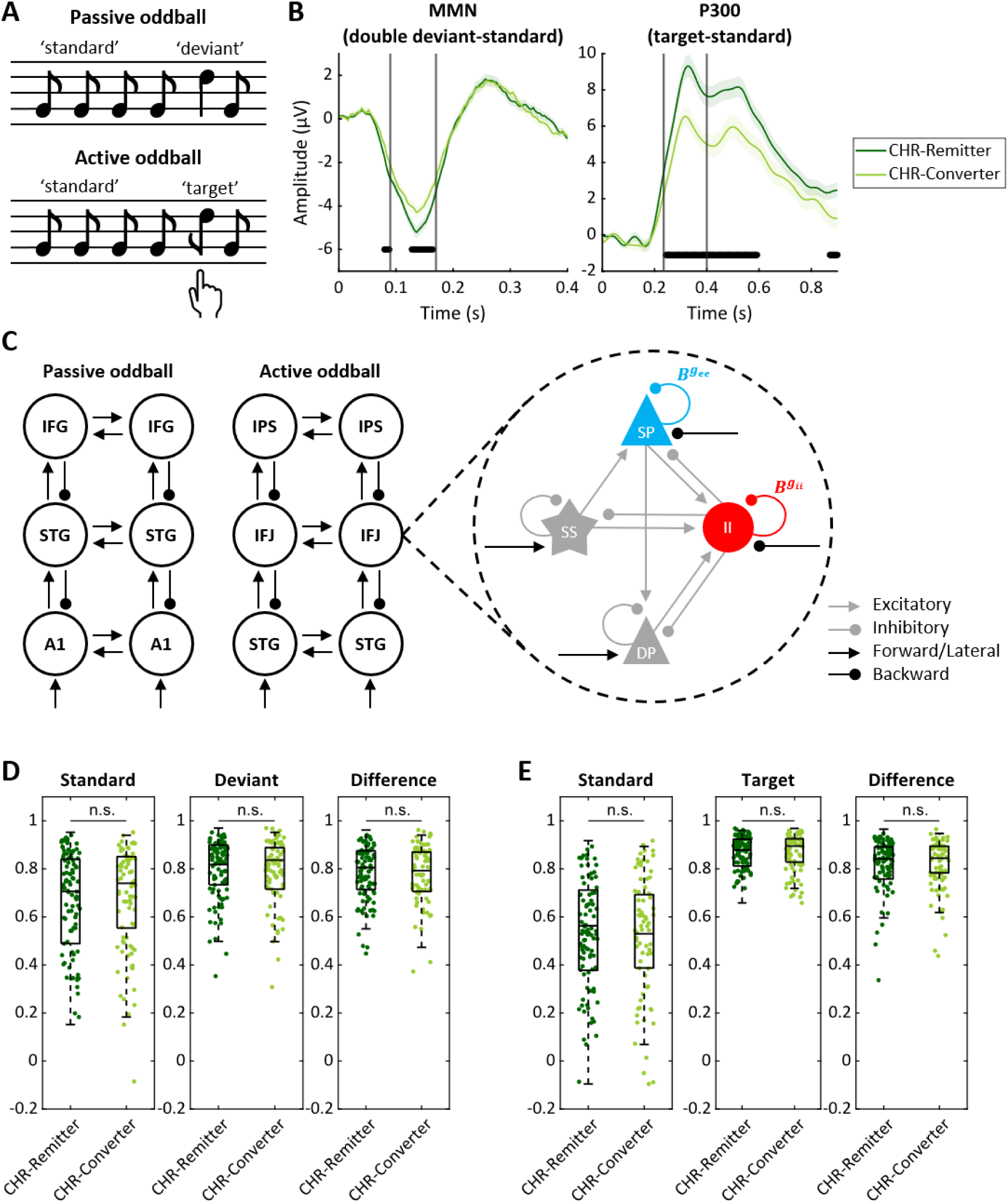
Dynamic causal modeling (DCM) of auditory oddball event-related potentials (ERP). **(A)** Simplified representations of the passive (top) and active (bottom) auditory oddball paradigms. Difference waves were computed by subtracting the ERP response to standard tones from the response to deviant or target tones. **(B)** Deviant-standard (left) and target-standard (right) ERP difference waves (mean ± SEM), plotted at 6 frontocentral electrodes (F3, Fz, F4, C3, Cz, C4) and at Pz, respectively, for participants at clinical high risk for psychosis who subsequently converted (CHR-Converter, light green) and those who remitted (CHR-Remitter, dark green). Vertical lines indicate the temporal windows used to measure mismatch negativity (MMN) and P300 components. Group differences in amplitude, computed using t-tests at each time point (uncorrected), are indicated below the difference waves. **(C)** Canonical microcircuit model for estimating E/I balance (53) used to analyze the two ERP paradigms. Bilateral sources in the primary auditory cortex (A1), superior temporal gyrus (STG) and inferior frontal gyrus (IFG) were used for the passive oddball paradigm. Bilateral sources in the STG, inferior frontal junction (IFJ) and intraparietal sulcus (IPS) were used for the active oddball paradigm. **(D)** Box and scatter plots showing the distribution of correlation coefficients across CHR-Converter and CHR-Remitter groups for standard tones (left), deviant tones (middle) and the deviant-standard difference wave (right). Each dot represents an individual participant. Group differences were tested using Mann-Whitney U tests applied to Fisher *z*-transformed correlation coefficients (n.s., not significant). Displayed values are untransformed for ease of interpretation. **(E)** Same as (D) for standard tones, target tones and the target-standard difference wave in the P300 analysis.

Additionally, *B*^*gee*^ and *B*^*gii*^ were constrained to be global (i.e., equal across regions). The motivation for this was two-fold. First, consistent with post-mortem (52,64–66), structural magnetic resonance imaging (67–69) findings, and with the polygenic nature of schizophrenia (47), we expected cellular pathology to be broadly distributed across cortical areas, rather than localized to a single region. Second, we limited the number of estimated parameters to reduce overfitting and obtain more efficient estimates of cortex-wide E/I balance. To further reduce the number of free parameters, we also imposed equality constraints on intrinsic connections and conduction delays, and fixed additional parameters (53). Priors over parameters are reported in **Table 2**, for inversion procedures see **Supplement**.

**Table 2.** Prior expectation and precision for each model parameter.

| Param. | Description | MMN<br>prior mean | P300<br>prior mean | Prior precision |
| --- | --- | --- | --- | --- |
| **Neuronal source model** |  |  |  |  |
| **A{1}** | Average extrinsic connectivity<br>(forward, SP to SS) | 0 | 0 | 1/16 |
| **A{2}** | Average extrinsic connectivity<br>(forward, SP to DP) | 0 | 0 | 1/16 |
| **A{3}** | Average extrinsic connectivity<br>(backward, DP to SP) | 0 | 0 | 1/16 |
| **A{4}** | Average extrinsic connectivity<br>(backward, DP to II) | 0 | 0 | 1/16 |
| **B** | Condition-specific extrinsic<br>connectivity | 0 | 0 | 1/8 |
| **C** | Input strength | [1 1 0 0 0 0] | [1 1 0 0 0 0] | [1/32 1/32 0 0 0 0] |
| **G** | Average intrinsic connectivity:<br>[$g_{ee}$ and $g_{ii}$] | 0 | 0 | 1/32 |
| **B<sup>g</sup>** | Condition-specific intrinsic<br>connectivity: [$B^{g_{ee}}$ and $B^{g_{ii}}$] | 0 | 0 | 1/8 |
| **T** | Synaptic time constant:<br>[SS SP II DP] | [16 32 2 128] ms | [32 128 2 2] ms | 1/32 |
| **D** | Neuronal delays (all sources):<br>[intrinsic extrinsic] | [1 8] ms | [1 8] ms | 1/64 |
| **S** | Slope of output firing rate<br>function | -1 | -1 | 1/64 |
| **R** | Stimulus: [onset dispersion] | [68 16] ms | [100 16] ms | 1/1024 |
| **Forward model** |  |  |  |  |
| **Lpos** | Source locations<br>[MNI coordinates] | [-42 -22 7;<br>46 -14 8;<br>-61 -32 8;<br>59 -25 8;<br>-46 20 8;<br>46 20 8] | [-61 -32 8;<br>59 -25 8;<br>-56 7 29;<br>50 8 30;<br>-33 -42 64;<br>33 -42 64] | 0 |
| **L** | Dipole orientation | 0 | 0 | 64 |
| **J** | Neuronal populations<br>contributing to the EEG:<br>[SS SP II DP] | [0 1 0 0] | [0 1 0 0] | 0 |

DP, deep pyramidal neurons; II, inhibitory interneurons; SP, superficial pyramidal neurons; SS, spiny stellate cells.

### Model diagnostics

Model fit (i.e., accuracy) was evaluated with the Pearson correlation (*r*) between observed and predicted responses to standard and deviant or target tones and the percentage variance explained (R²). Correlation coefficients were transformed using Fisher’s *r*-to-*z* transformation for statistical testing (**Figures S1A and S1B**). Due to non-normal distributions indicated by Kolmogorov-Smirnov tests, group differences in Fisher *z*-transformed *r* and R² values were examined using Mann-Whitney U tests.

To evaluate parameter identifiability, we computed Pearson correlations between *B*^*gee*^ and *B*^*gii*^. High correlations (r>|0.6|) (70) between parameters can reflect the model’s inability to disentangle their distinct contributions; in contrast, low correlations suggest that the model is able to estimate each parameter’s unique effect; i.e., disentangle or resolve conditional dependencies.

### Second-level analysis

We used parametric empirical Bayes (PEB) (71,72) to test for second-level (between-subject) effects on synaptic parameters. PEB is a Bayesian general linear model that takes both the expectation and covariance of each participant’s individual (first-level) parameter estimate into account. We explain variance in first-level E/I parameters (*B*^*gee*^ and *B*^*gii*^) across participants with variables of interest (group or symptom severity) and covariates (age, study site, antipsychotic medication). Age-by-group interactions were tested and excluded from the PEB analysis due to lack of significance (**Figure S2**). We report the Bayesian posterior probability (P) for each effect, which quantifies the probability that the effect is present (i.e., non-zero), given the data and model. A P>0.95 is interpreted as strong evidence for an effect (73).

### Posterior predictive checks

To explore the contribution of *B*^*gee*^ and *B*^*gii*^, we conducted simulations using posterior parameter estimates from the grand mean inversion (**Supplement**). In each simulation, a single parameter was systematically varied while all the other parameters were held constant at their posterior to evaluate its effect on ERP amplitude. This (sensitivity) analysis discloses the contribution of each parameter to the data features being explained; in this instance the MMN and P300.

## Results

### CHR-Converters show reduced MMN and P300 amplitudes compared to CHR-Remitters

We analyzed baseline ERP data from passive and active auditory oddball paradigms (**Figure 1A**) in CHR-P participants who converted to psychosis within two years and those who remitted. MMN amplitude (90-170 ms (24)) was significantly reduced in CHR-Converters compared to CHR-Remitters (t_169_=-2.22, p=.028). Timepoint-wise t-tests (uncorrected) identified the most prominent effects around 130-160 ms post-stimulus (**Figure 1B, left**). Amplitude of the P3b subcomponent (235-400 ms (27)) was also reduced in CHR-Converters compared to CHR-Remitters (t_160_=-3.22, p=.002). Timepoint-wise t-tests (uncorrected) showed that this deficit extended to the slow wave following the P3b peak (400-630 ms; **Figure 1B, right**).

### Excitation/inhibition model captures individual variability in MMN and P300

Our computational model (**Figure 1C**) adequately fit most participants’ MMN (**Figure 1D**) and P300 (**Figure 1E**) data, with mean (SD) correlations between observed and predicted responses of *r*=0.78 (0.12) for the deviant-standard difference waveform and *r*=0.81 (0.11) for the target-standard difference waveform. Mean R² was 0.61 (0.18) for the deviant-standard and 0.67 (0.16) for the target-standard difference waveform. Model fit did not differ significantly between groups (all p>0.05).

Correlations between parameter estimates across subjects were unconcerning (r=-0.04 for MMN, r=0.20 for P300; **Figures S1C and S1D**), indicating good parameter identifiability.

### Computational modeling indicates a loss of pyramidal cell excitability in CHR-Converters

Across both paradigms, we found a consistent condition-specific increase in pyramidal self-inhibition (i.e., decreased excitability) in deviant relative to standard trials (P>.99) and target relative to standard trials (P>.95) in CHR-Converters compared to CHR-Remitters (**Figures 2A and 2B**). The evidence for these effects remained strong when omitting age and site covariates, and robust to the inclusion of antipsychotic-use covariates (**Figures S4A and S4B**). In the unmedicated subsample (n=54 CHR-Converters, n=78 CHR-Remitters), the group difference in pyramidal self-inhibition remained strong for the active (P>.95; **Figure S4D**) but not the passive (P=.68; **Figure S4C**) oddball.

**Figure 2.**
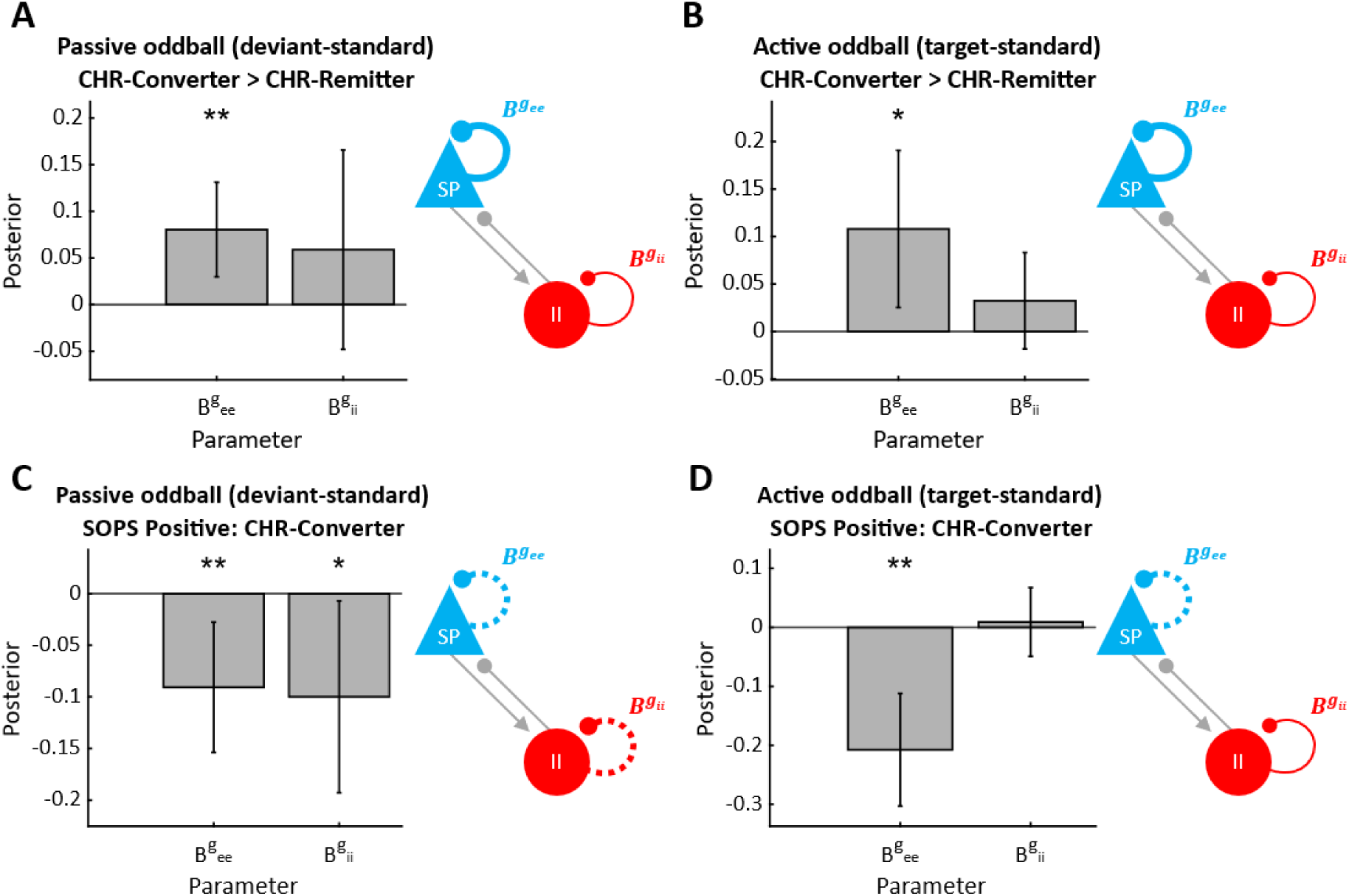
Group-level analysis of mismatch negativity (MMN) and P300 model parameters. Panels. **(A)** and **(B)** show group differences in condition-specific modulation of superficial pyramidal cell self-inhibition (*B*^*gee*^) and inhibitory interneuron self-inhibition (*B*^*gii*^) in clinical high risk for psychosis (CHR-P) participants who subsequently converted to psychosis (CHR-Converter) compared to CHR-P participants whose symptoms remitted at follow-up (CHR-Remitter) for **(A)** the MMN paradigm and **(B)** the P300 paradigm. Asterisks (*) indicate a significant group effect with a posterior probability of P>.95. In the microcircuit diagram, parameter increases are illustrated with thick lines. Panels **(C)** and **(D)** show condition-specific microcircuit parameters associated with Scale of Psychosis-Risk Syndromes (SOPS) positive symptoms in **(C)** the MMN paradigm and **(D)** the P300 paradigm. Asterisks indicate a significant correlation with a posterior probability of (*) P>.95 or (**) P>.99. In the microcircuit diagram, negative associations are illustrated with dotted lines. Across all panels, error bars represent the 95% Bayesian confidence intervals.

### Positive symptoms are associated with increased, rather than decreased, pyramidal cell excitability

Across CHR-Converters and CHR-Remitters, there were no associations between SOPS Positive scores and pyramidal or interneuron excitability. However, within CHR-Converters, more severe baseline positive symptoms were associated with decreased pyramidal self-inhibition (i.e., disinhibition) across both tasks (MMN: P>.95, P300: P>.99; **Figures 2C and 2D**), and decreased self-inhibition of interneurons during the MMN paradigm (P>.95; **Figure 2C**).

### Posterior predictive checks confirm that reduced pyramidal cell excitability explains ERP deficits

Across both paradigms, reducing pyramidal excitability in simulations resulted in reduced MMN (**Figure 3A, top**) and P300 (**Figure 3B, top**) amplitudes – as observed empirically in CHR-Converters. This reduction was caused by a strong suppression of the deviant/target ERP and weakly increased standard ERPs. In contrast, increased interneuron self-inhibition (i.e., reduced interneuron excitability) had the opposite effect, resulting in larger – not smaller – MMN (**Figure 3A, bottom**) and P300 (**Figure 3B, bottom**) amplitudes. These simulations suggest that reduced pyramidal, but not interneuron excitability, can explain the empirical effects in CHR-Converters.

**Figure 3.**
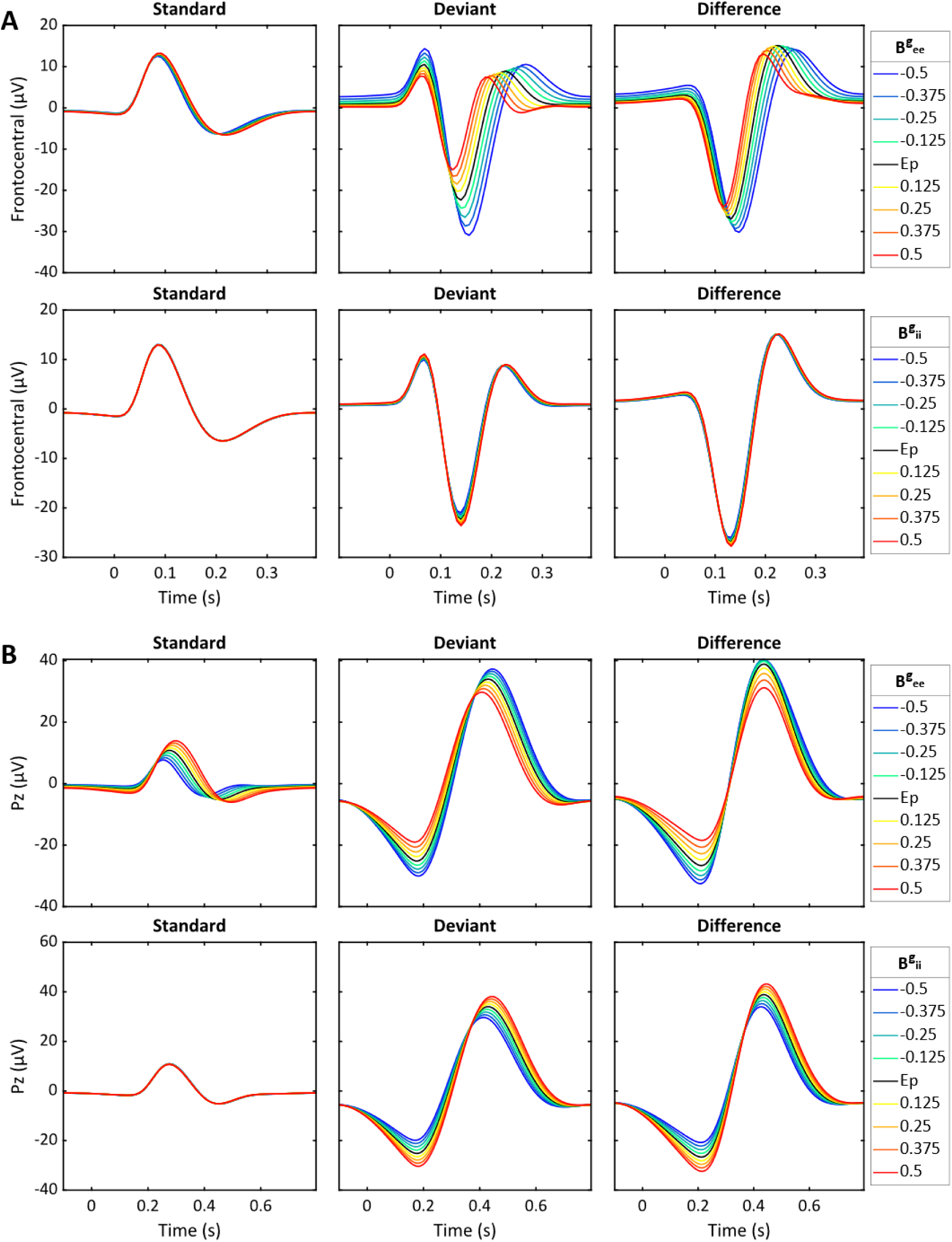
Microcircuit parameter sensitivity analyses. This figure shows simulations derived from the grand mean model illustrating the effects of varying superficial pyramidal cell self-inhibition (*B*^*gee*^, top) and inhibitory interneuron self-inhibition (*B*^*gii*^, bottom) on **(A)** the deviant-standard difference waveform at 6 frontocentral electrodes (F3, Fz, F4, C3, Cz, C4) and **(B)** the target-standard difference waveform at electrode Pz. Each line represents a different simulation, in which the connectivity strength of the self-connection was scaled by ±50% around the posterior estimate (Ep), which is shown in black. Warmer colors indicate parameter increases, and cooler colors represent reductions.

Note that the E/I parameters model the difference between conditions and thus have opposite effects on the standard and deviant/target by design. However, here, we assessed – through simulations – which synaptic effect could explain MMN and P300 reductions.

### Age and antipsychotic use affect pyramidal cell excitability during the MMN

Across both groups, older age was associated with decreased pyramidal self-inhibition (i.e., disinhibition) during the MMN (P>.99; **Figure S2A**), but not the P300 paradigm (**Figure S2B**). There was no evidence of age-by-group interactions in either paradigm (**Figures S2C and S2D**).

There was no main effect of antipsychotic use in either paradigm (**Figures S3A and S3B**). However, an antipsychotic use-by-group interaction was observed for pyramidal excitability during the MMN (P>.95; **Figure S3C**), but not the P300 paradigm (**Figure S3D**). Follow-up tests indicated that antipsychotic use was associated with increased pyramidal self-inhibition (i.e., decreased excitability) in CHR-Converters (P>.99; **Figure S3E**) but decreased pyramidal self-inhibition (i.e., disinhibition) in CHR-Remitters (P>.95; **Figure S3F**).

## Discussion

In what we believe to be the largest longitudinal study of CHR-P individuals to date, previously established ERP deficits in CHR-Converters compared to CHR-Remitters – across two auditory oddball paradigms (24,27) – were best explained by a reduction in pyramidal cell excitability in future CHR-Converters. For both passive (MMN) and active (target P300) oddball paradigms, baseline positive symptom severity within CHR-Converters was associated with the opposite circuit change: pyramidal cell disinhibition. Together, these findings support the hypothesis that, in the early stages of psychotic illness, a primary loss of pyramidal cell excitability is compensated by an allostatic downregulation of inhibition (32,44) – thereby restoring E/I balance. Under this hypothesis, secondary downregulation precipitates psychotic symptoms.

Crucially, our findings provide evidence that a loss of pyramidal cell excitability – previously observed in chronic schizophrenia (51) and, more recently, in two of three transdiagnostic biotypes composed of people with schizophrenia, bipolar, and schizoaffective disorder (74) – is already present in youth at CHR-P who develop psychosis within 2 years. We observed this deficit in CHR-Converters compared to CHR-Remitters, suggesting that excitatory dysfunction is not merely a general feature of psychosis risk but a potential mechanistic marker underlying progression from the CHR-P syndrome to full psychosis. This also supports the idea that excitatory dysfunction is fundamental to schizophrenia, rather than just a consequence of chronic psychotic illness.

Although 17% of CHR-Remitters and 30% of CHR-Converters were taking antipsychotic medications at baseline, our results were unaltered when antipsychotic use was included as a covariate (cf. (51)), indicating that the observed deficits were unlikely to be driven by medication effects. Analyses restricted to unmedicated participants showed pyramidal cell hypoexcitability in CHR-Converters compared to CHR-Remitters during the active, but not the passive, oddball task; possibly due to a loss of power or because this subsample likely underrepresents more severe cases (75–77), introducing a potential confound. Additional analyses also revealed an interaction between group and antipsychotic use on pyramidal cell excitability during the passive oddball.

Reduced condition-specific pyramidal cell excitability in CHR-Converters may reflect hypofunction of NMDA receptors and their signaling cascade – and a disproportionate effect of this on the larger oddball waveforms – as NMDA receptor antagonism has consistently been shown to reduce MMN (35,36,38,78,79) and P300 (36,40–42) amplitudes. While this change in synaptic efficacy could also be attributed to increased inhibition via parvalbumin-positive (PV) interneurons, postmortem studies suggest that PV interneurons are *hypo*active, rather than hyperactive, in schizophrenia (80–82). We did not find evidence of such a reduction in PV function in CHR-Converters. Our findings challenge the hypothesis that interneuron abnormalities are a primary mechanism in schizophrenia and point to pyramidal cell NMDA receptor hypofunction as a more plausible explanation. This is consistent with recent work suggesting that PV interneuron deficits are secondary to reduced excitatory drive (31,44).

While reduced pyramidal cell excitability distinguished CHR-Converters from CHR-Remitters, a somewhat paradoxical finding was that greater symptom severity was associated with disinhibition in both paradigms. This opposing effect mirrors findings from a previous study in chronic schizophrenia: across EEG and fMRI paradigms, people with schizophrenia showed reduced pyramidal cell excitability compared to controls, but positive (auditory) symptoms correlated with disinhibition in auditory areas (51). At the sensor-level, 40 Hz power and phase-locking in the auditory steady state paradigm are both reduced in schizophrenia, yet they correlate positively with auditory symptoms (83,84). Similarly, a mega-analysis of magnetic resonance spectroscopy studies found generally reduced levels of glutamate in people with schizophrenia, but a positive correlation between glutamate and positive symptoms (85). These findings suggest that it is compensatory disinhibition, and not the primary glutamatergic dysfunction, that leads to psychotic symptoms.

Our study is not without limitations. First, the design limits our ability to draw causal inferences regarding the role of E/I (im)balance. Although we stratified CHR-P participants by clinical outcome, longitudinal studies are needed to track E/I balance across illness stages; particularly in genetically at-risk individuals, where E/I balance could be studied before the onset of prodromal symptoms. While 1-year and 2-year EEG follow-up data were available in NAPLS2, most conversions occurred within the first year, limiting their utility for understanding pre-conversion changes. Studies with shorter follow-up intervals may help address this (86). Longitudinal designs will also be key for establishing the test-retest reliability connectivity or synaptic efficacy estimates, for instance by assessing the impact of measurement error and state vs trait-like effects (87).

Another key consideration is the potential impact of ‘accelerated aging’ in schizophrenia (88–93), which has been shown to affect E/I balance (94,95). Notably, we saw that older age was associated with disinhibition during the MMN paradigm, but found no evidence of age-by-group interactions in pyramidal or interneuron excitability, meaning the observed changes in future CHR-Converters were unlikely to be driven by accelerated aging. As a result, we included only the main effect of age in second-level models. This contrasts with prior analyses of NAPLS2 data, which incorporated the group-by-age interaction into the disease effect, effectively adjusting for ‘healthy aging’ (24,27).

In conclusion, biophysical modeling of two auditory oddball paradigms revealed two key findings: a consistent reduction in superficial pyramidal cell excitability in CHR-Converters compared to CHR-Remitters, and an association between positive symptom severity and disinhibition in CHR-Converters. This suggests that reduced pyramidal excitability is a primary aetiological factor in CHR-Converters (as it is in schizophrenia). Conversely, disinhibition in cortical circuits may be a compensatory response to pyramidal cell dysfunction contributing to the emergence of psychotic symptoms.

## Code availability

The analysis code will be made publicly available at https://github.com/juliarodsan/napls_ei upon acceptance of the paper. The model code will be made publicly available at https://github.com/daniel-hauke/dcm_ei (Hauke et al., 2025) upon acceptance of the paper.

## Data availability

Access to NAPLS2 EEG data can be arranged through a formal collaboration with the NAPLS2 PIs. Contact Dr. Daniel Mathalon to inquire about such collaborations.

## Competing Interests

The authors declare no competing interests.

## Funding

This work was supported by the National Institute of Mental Health (grants U01MH081902 to Dr. Cannon, P50 MH066286 to Dr. Bearden, U01MH081988 to Dr. Walker, U01MH076989 to Dr. Mathalon, U01MH081944 to Dr. Cadenhead, U01MH081984 to Dr. Addington, U01MH082004 to Dr. Perkins, U01MH081857 to Dr. Cornblatt, U01MH081928 to Dr. Seidman, and U01MH082022 to Dr. Woods) and the Medical Research Council (MR/N013867/1 to Dr. Rodriguez-Sanchez). Drs. Rodriguez-Sanchez, Hauke, Pinotsis and Adams are funded by Dr. Adams’ Future Leaders Fellowship (MR/W011751/1). Prof. Friston is supported by funding from the Wellcome Trust (226793/Z/22/Z).

Drs. Hamilton, Mathalon, Duncan, Light, and Niznikiewicz are employees of the US government. The content is solely the responsibility of the authors and does not necessarily reflect the position or policy of the National Institutes of Health, the Department of Veterans Affairs, or the US government.

## Acknowledgements

We thank Drs. Larry J. Seidman, PhD and Barbara A. Cornblatt, who passed away before this manuscript was submitted, for their contributions to this work.

We thank Drs. Thomas H. McGlashan and Ming T. Tsuang, who were not available to review the current manuscript, for their contributions to this work.

## Notes

### Competing Interest Statement

The authors have declared no competing interest.

### Funding Statement

This study was funded by the National Institute of Mental Health (grants U01MH081902 to Dr. Cannon, P50 MH066286 to Dr. Bearden, U01MH081988 to Dr. Walker, U01MH076989 to Dr. Mathalon, U01MH081944 to Dr. Cadenhead, U01MH081984 to Dr. Addington, U01MH082004 to Dr. Perkins, U01MH081857 to Dr. Cornblatt, U01MH081928 to Dr. Seidman, and U01MH082022 to Dr. Woods) and the Medical Research Council (MR/N013867/1 to Dr. Rodriguez-Sanchez). Drs. Hauke, Rodriguez-Sanchez, Pinotsis and Adams are funded by Dr. Adams' Future Leaders Fellowship (MR/W011751/1). Dr. Friston is supported by funding from the Wellcome Trust (226793/Z/22/Z).

### Author Declarations

The Institutional Review Boards of Emory University, Harvard Medical School, University of Calgary, University of California, Los Angeles, University of California, San Diego, University of North Carolina at Chapel Hill, Yale University, and Zucker Hillside Hospital gave ethical approval for this work.

## References

1. McGlashan T, Walsh B, Woods S (2010): The Psychosis-Risk Syndrome: Handbook for Diagnosis and Follow-Up. Oxford University Press.

2. Inns M, Gill M, Arribas M, Logeswaran Y, Minichino A, Reilly TJ, et al. (2025, June 26): Can we detect the undetected? Comparing the prodromes of individuals with first episode psychosis detected and undetected by clinical high risk for psychosis services: an electronic health record study. MedRxiv. 10.1101/2025.06.26.25330305

3. Fusar-Poli P, Cappucciati M, Borgwardt S, Woods SW, Addington J, Nelson B, et al. (2016): Heterogeneity of Psychosis Risk Within Individuals at Clinical High Risk: A Meta-analytical Stratification. JAMA Psychiatry 73: 113–120.

4. Fusar-Poli P (2017): The Clinical High-Risk State for Psychosis (CHR-P), Version II. Schizophr Bull 43: 44–47.

5. Perkins DO, Jeffries CD, Cornblatt BA, Woods SW, Addington J, Bearden CE, et al. (2015): Severity of thought disorder predicts psychosis in persons at clinical high-risk. Schizophr Res 169: 169–177.

6. Simon AE, Borgwardt S, Riecher-Rössler A, Velthorst E, de Haan L, Fusar-Poli P (2013): Moving beyond transition outcomes: meta-analysis of remission rates in individuals at high clinical risk for psychosis. Psychiatry Res 209: 266–272.

7. Davies C, Cipriani A, Ioannidis JPA, Radua J, Stahl D, Provenzani U, et al. (2018): Lack of evidence to favor specific preventive interventions in psychosis: a network meta-analysis. World Psychiatry 17: 196–209.

8. Näätänen R, Teder W, Alho K, Lavikainen J (1992): Auditory attention and selective input modulation: a topographical ERP study. Neuroreport 3: 493–496.

9. Braff DL, Light GA (2004): Preattentional and attentional cognitive deficits as targets for treating schizophrenia. Psychopharmacology (Berl) 174: 75–85.

10. Garrido MI, Kilner JM, Stephan KE, Friston KJ (2009): The mismatch negativity: a review of underlying mechanisms. Clin Neurophysiol 120: 453–463.

11. Lieder F, Daunizeau J, Garrido MI, Friston KJ, Stephan KE (2013): Modelling trial-by-trial changes in the mismatch negativity. PLoS Comput Biol 9: e1002911.

12. Näätänen R, Jacobsen T, Winkler I (2005): Memory-based or afferent processes in mismatch negativity (MMN): a review of the evidence. Psychophysiology 42: 25–32.

13. Todd J, Michie PT, Schall U, Ward PB, Catts SV (2012): Mismatch negativity (MMN) reduction in schizophrenia-impaired prediction--error generation, estimation or salience? Int J Psychophysiol 83: 222–231.

14. Polich J (2007): Updating P300: an integrative theory of P3a and P3b. Clin Neurophysiol 118: 2128–2148.

15. Debener S, Kranczioch C, Herrmann CS, Engel AK (2002): Auditory novelty oddball allows reliable distinction of top-down and bottom-up processes of attention. Int J Psychophysiol 46: 77–84.

16. Erickson MA, Ruffle A, Gold JM (2016): A Meta-Analysis of Mismatch Negativity in Schizophrenia: From Clinical Risk to Disease Specificity and Progression. Biol Psychiatry 79: 980–987.

17. Umbricht D, Krljes S (2005): Mismatch negativity in schizophrenia: a meta-analysis. Schizophr Res 76: 1–23.

18. Bramon E, Rabe-Hesketh S, Sham P, Murray RM, Frangou S (2004): Meta-analysis of the P300 and P50 waveforms in schizophrenia. Schizophr Res 70: 315–329.

19. Jeon Y-W, Polich J (2003): Meta-analysis of P300 and schizophrenia: patients, paradigms, and practical implications. Psychophysiology 40: 684–701.

20. Hamilton HK, Boos AK, Mathalon DH (2020): Electroencephalography and event-related potential biomarkers in individuals at clinical high risk for psychosis. Biol Psychiatry 88: 294–303.

21. Hamilton HK, Mathalon DH (2024): Neurophysiological models in individuals at clinical high risk for psychosis: Using translational EEG paradigms to forecast psychosis risk and resilience. Adv Neurobiol 40: 385–410.

22. Bodatsch M, Ruhrmann S, Wagner M, Müller R, Schultze-Lutter F, Frommann I, et al. (2011): Prediction of psychosis by mismatch negativity. Biol Psychiatry 69: 959–966.

23. Fryer SL, Roach BJ, Hamilton HK, Bachman P, Belger A, Carrión RE, et al. (2020): Deficits in auditory predictive coding in individuals with the psychosis risk syndrome: Prediction of conversion to psychosis. J Abnorm Psychol 129: 599–611.

24. Hamilton HK, Roach BJ, Bachman PM, Belger A, Carrión RE, Duncan E, et al. (2022): Mismatch Negativity in Response to Auditory Deviance and Risk for Future Psychosis in Youth at Clinical High Risk for Psychosis. JAMA Psychiatry 79: 780–789.

25. Kim M, Lee TH, Yoon YB, Lee TY, Kwon JS (2018): Predicting remission in subjects at clinical high risk for psychosis using mismatch negativity. Schizophr Bull 44: 575–583.

26. Perez VB, Woods SW, Roach BJ, Ford JM, McGlashan TH, Srihari VH, Mathalon DH (2014): Automatic auditory processing deficits in schizophrenia and clinical high-risk patients: forecasting psychosis risk with mismatch negativity. Biol Psychiatry 75: 459–469.

27. Hamilton HK, Roach BJ, Bachman PM, Belger A, Carrion RE, Duncan E, et al. (2019): Association Between P300 Responses to Auditory Oddball Stimuli and Clinical Outcomes in the Psychosis Risk Syndrome. JAMA Psychiatry 76: 1187–1197.

28. Hamilton HK, Woods SW, Roach BJ, Llerena K, McGlashan TH, Srihari VH, et al. (2019): Auditory and visual oddball stimulus processing deficits in schizophrenia and the psychosis risk syndrome: Forecasting psychosis risk with P300. Schizophr Bull 45: 1068–1080.

29. van Tricht MJ, Nieman DH, Koelman JHTM, van der Meer JN, Bour LJ, de Haan L, Linszen DH (2010): Reduced parietal P300 amplitude is associated with an increased risk for a first psychotic episode. Biol Psychiatry 68: 642–648.

30. Kim M, Lee TY, Lee S, Kim SN, Kwon JS (2015): Auditory P300 as a predictor of short-term prognosis in subjects at clinical high risk for psychosis. Schizophr Res 165: 138–144.

31. Dienel SJ, Lewis DA (2019): Alterations in cortical interneurons and cognitive function in schizophrenia. Neurobiol Dis 131: 104208.

32. Krystal JH, Anticevic A, Yang GJ, Dragoi G, Driesen NR, Wang X-J, Murray JD (2017): Impaired Tuning of Neural Ensembles and the Pathophysiology of Schizophrenia: A Translational and Computational Neuroscience Perspective. Biol Psychiatry 81: 874–885.

33. Díez Á, Ranlund S, Pinotsis D, Calafato S, Shaikh M, Hall M-H, et al. (2017): Abnormal frontoparietal synaptic gain mediating the P300 in patients with psychotic disorder and their unaffected relatives. Hum Brain Mapp 38: 3262–3276.

34. Ranlund S, Adams RA, Díez Á, Constante M, Dutt A, Hall M-H, et al. (2016): Impaired prefrontal synaptic gain in people with psychosis and their relatives during the mismatch negativity. Hum Brain Mapp 37: 351–365.

35. Javitt DC, Steinschneider M, Schroeder CE, Arezzo JC (1996): Role of cortical N-methyl-D-aspartate receptors in auditory sensory memory and mismatch negativity generation: implications for schizophrenia. Proc Natl Acad Sci U S A 93: 11962–11967.

36. Gunduz-Bruce H, Reinhart RMG, Roach BJ, Gueorguieva R, Oliver S, D’Souza DC, et al. (2012): Glutamatergic modulation of auditory information processing in the human brain. Biol Psychiatry 71: 969–977.

37. Schwertner A, Zortea M, Torres FV, Caumo W (2018): Effects of subanesthetic ketamine administration on visual and auditory event-related potentials (ERP) in humans: A systematic review. Front Behav Neurosci 12: 70.

38. Umbricht D, Schmid L, Koller R, Vollenweider FX, Hell D, Javitt DC (2000): Ketamine-induced deficits in auditory and visual context-dependent processing in healthy volunteers: implications for models of cognitive deficits in schizophrenia. Arch Gen Psychiatry 57: 1139– 1147.

39. Rosburg T, Kreitschmann-Andermahr I (2016): The effects of ketamine on the mismatch negativity (MMN) in humans - A meta-analysis. Clin Neurophysiol 127: 1387–1394.

40. Mathalon DH, Ahn K-H, Perry EB Jr, Cho H-S, Roach BJ, Blais RK, et al. (2014): Effects of nicotine on the neurophysiological and behavioral effects of ketamine in humans. Front Psychiatry 5: 3.

41. Oranje B, van Berckel BN, Kemner C, van Ree JM, Kahn RS, Verbaten MN (2000): The effects of a sub-anaesthetic dose of ketamine on human selective attention. Neuropsychopharmacology 22: 293–302.

42. Watson TD, Petrakis IL, Edgecombe J, Perrino A, Krystal JH, Mathalon DH (2009): Modulation of the cortical processing of novel and target stimuli by drugs affecting glutamate and GABA neurotransmission. Int J Neuropsychopharmacol 12: 357–370.

43. Rowland LM, Summerfelt A, Wijtenburg SA, Du X, Chiappelli JJ, Krishna N, et al. (2016): Frontal Glutamate and γ-Aminobutyric Acid Levels and Their Associations With Mismatch Negativity and Digit Sequencing Task Performance in Schizophrenia. JAMA Psychiatry 73: 166–174.

44. Chung DW, Fish KN, Lewis DA (2016): Pathological Basis for Deficient Excitatory Drive to Cortical Parvalbumin Interneurons in Schizophrenia. Am J Psychiatry 173: 1131–1139.

45. Glantz LA, Lewis DA (2000): Decreased dendritic spine density on prefrontal cortical pyramidal neurons in schizophrenia. Arch Gen Psychiatry 57: 65–73.

46. Glausier JR, Lewis DA (2013): Dendritic spine pathology in schizophrenia. Neuroscience 251: 90– 107.

47. Trubetskoy V, Pardiñas AF, Qi T, Panagiotaropoulou G, Awasthi S, Bigdeli TB, et al. (2022): Mapping genomic loci implicates genes and synaptic biology in schizophrenia. Nature 604: 502–508.

48. Gao R, Peterson EJ, Voytek B (2017): Inferring synaptic excitation/inhibition balance from field potentials. Neuroimage 158: 70–78.

49. Diehl GW, Redish AD (2024, January 24): Measuring excitation-inhibition balance through spectral components of local field potentials. BioRxivorg. 10.1101/2024.01.24.577086

50. Stephan KE, Schlagenhauf F, Huys QJM, Raman S, Aponte EA, Brodersen KH, et al. (2017): Computational neuroimaging strategies for single patient predictions. Neuroimage 145: 180–199.

51. Adams RA, Pinotsis D, Tsirlis K, Unruh L, Mahajan A, Horas AM, et al. (2022): Computational Modeling of Electroencephalography and Functional Magnetic Resonance Imaging Paradigms Indicates a Consistent Loss of Pyramidal Cell Synaptic Gain in Schizophrenia. Biol Psychiatry 91: 202–215.

52. Osimo EF, Beck K, Reis Marques T, Howes OD (2019): Synaptic loss in schizophrenia: a meta-analysis and systematic review of synaptic protein and mRNA measures. Mol Psychiatry 24: 549–561.

53. Hauke DJ, Rodriguez-Sanchez J, Oloye H, Berndt L, Pinotsis D, Friston KJ, et al. (2025): A canonical microcircuit for estimating excitation/inhibition (E/I) balance.

54. Addington J, Cadenhead KS, Cornblatt BA, Mathalon DH, McGlashan TH, Perkins DO, et al. (2012): North American Prodrome Longitudinal Study (NAPLS 2): overview and recruitment. Schizophr Res 142: 77–82.

55. Addington J, Liu L, Buchy L, Cadenhead KS, Cannon TD, Cornblatt BA, et al. (2015): North American Prodrome Longitudinal Study (NAPLS 2): The Prodromal Symptoms. J Nerv Ment Dis 203: 328–335.

56. Schöbi D (2020): Dynamic Causal Models for Inference on Neuromodulatory Processes in Neural Circuits. ETH Zurich. 10.3929/ETHZ-B-000429311

57. Doeller CF, Opitz B, Mecklinger A, Krick C, Reith W, Schröger E (2003): Prefrontal cortex involvement in preattentive auditory deviance detection: neuroimaging and electrophysiological evidence. NeuroImage 20: 1270–1282.

58. Molholm S, Martinez A, Ritter W, Javitt DC, Foxe JJ (2005): The neural circuitry of pre-attentive auditory change-detection: an fMRI study of pitch and duration mismatch negativity generators. Cereb Cortex 15: 545–551.

59. Opitz B, Rinne T, Mecklinger A, von Cramon DY, Schröger E (2002): Differential contribution of frontal and temporal cortices to auditory change detection: fMRI and ERP results. Neuroimage 15: 167–174.

60. Garrido MI, Friston KJ, Kiebel SJ, Stephan KE, Baldeweg T, Kilner JM (2008): The functional anatomy of the MMN: a DCM study of the roving paradigm. Neuroimage 42: 936–944.

61. Kim H (2014): Involvement of the dorsal and ventral attention networks in oddball stimulus processing: a meta-analysis. Hum Brain Mapp 35: 2265–2284.

62. Bastos AM, Usrey WM, Adams RA, Mangun GR, Fries P, Friston KJ (2012): Canonical microcircuits for predictive coding. Neuron 76: 695–711.

63. Shaw AD, Moran RJ, Muthukumaraswamy SD, Brealy J, Linden DE, Friston KJ, Singh KD (2017): Neurophysiologically-informed markers of individual variability and pharmacological manipulation of human cortical gamma. Neuroimage 161: 19–31.

64. Fish KN, Rocco BR, DeDionisio AM, Dienel SJ, Sweet RA, Lewis DA (2021): Altered Parvalbumin Basket Cell Terminals in the Cortical Visuospatial Working Memory Network in Schizophrenia. Biol Psychiatry 90: 47–57.

65. Selemon LD, Rajkowska G, Goldman-Rakic PS (1995): Abnormally high neuronal density in the schizophrenic cortex. A morphometric analysis of prefrontal area 9 and occipital area 17. Arch Gen Psychiatry 52: 805–18; discussion 819-20.

66. Selemon LD, Rajkowska G, Goldman-Rakic PS (1998): Elevated neuronal density in prefrontal area 46 in brains from schizophrenic patients: Application of a three-dimensional, stereologic counting method. J Comp Neurol 392: 402–412.

67. Zipursky RB, Lim KO, Sullivan EV, Brown BW, Pfefferbaum A (1992): Widespread cerebral gray matter volume deficits in schizophrenia. Arch Gen Psychiatry 49: 195–205.

68. Lim KO, Sullivan EV, Zipursky RB, Pfefferbaum A (1996): Cortical gray matter volume deficits in schizophrenia: a replication. Schizophr Res 20: 157–164.

69. Shenton ME, Dickey CC, Frumin M, McCarley RW (2001): A review of MRI findings in schizophrenia. Schizophr Res 49: 1–52.

70. Hauke DJ, Wobmann M, Andreou C, Mackintosh AJ, de Bock R, Karvelis P, et al. (2024): Altered perception of environmental volatility during social learning in emerging psychosis. Comput Psychiatr 8: 1–22.

71. Friston KJ, Litvak V, Oswal A, Razi A, Stephan KE, van Wijk BCM, et al. (2016): Bayesian model reduction and empirical Bayes for group (DCM) studies. Neuroimage 128: 413–431.

72. Zeidman P, Jafarian A, Seghier ML, Litvak V, Cagnan H, Price CJ, Friston KJ (2019): A guide to group effective connectivity analysis, part 2: Second level analysis with PEB. Neuroimage 200: 12–25.

73. Kass RE, Raftery AE (1995): Bayes Factors. J Am Stat Assoc 90: 773.

74. Oloye H, Hauke DJ, Rodriguez-Sanchez J, Parker D, Pearlson G, Keshavan M, et al. (2025): Computational modelling of paired click and auditory oddball ERP responses to probe inhibitory and excitatory cell function in schizophrenia and across transdiagnostic biotypes: Insights from the B-SNIP study. In preparation.

75. Mukhtar H, Zhou D, Farina EA, Saxena A, Cahill J, Addington J, et al. (2025): Prediction of antipsychotic medication inception in antipsychotic-naive youth at clinical high risk for psychosis. Psychol Med 55. 10.1017/s0033291725101372

76. Zeng J, Raballo A, Gan R, Wu G, Wei Y, Xu L, et al. (2022): Antipsychotic exposure in clinical high risk of psychosis: Empirical insights from a large cohort study. J Clin Psychiatry 83. 10.4088/JCP.21m14092

77. Pelizza L, Leuci E, Quattrone E, Azzali S, Paulillo G, Pupo S, et al. (2024): Baseline antipsychotic prescription and short-term outcome indicators in individuals at clinical high-risk for psychosis: Findings from the Parma At-Risk Mental States (PARMS) program. Early Interv Psychiatry 18: 71–81.

78. Todd J, Harms L, Schall U, Michie PT (2013): Mismatch negativity: translating the potential. Front Psychiatry 4: 171.

79. Hamilton HK, D’Souza DC, Ford JM, Roach BJ, Kort NS, Ahn K-H, et al. (2018): Interactive effects of an N-methyl-d-aspartate receptor antagonist and a nicotinic acetylcholine receptor agonist on mismatch negativity: Implications for schizophrenia. Schizophr Res 191: 87–94.

80. Curley AA, Arion D, Volk DW, Asafu-Adjei JK, Sampson AR, Fish KN, Lewis DA (2011): Cortical deficits of glutamic acid decarboxylase 67 expression in schizophrenia: clinical, protein, and cell type-specific features. Am J Psychiatry 168: 921–929.

81. Gonzalez-Burgos G, Hashimoto T, Lewis DA (2010): Alterations of cortical GABA neurons and network oscillations in schizophrenia. Curr Psychiatry Rep 12: 335–344.

82. Guidotti A, Auta J, Davis JM, Di-Giorgi-Gerevini V, Dwivedi Y, Grayson DR, et al. (2000): Decrease in reelin and glutamic acid decarboxylase67 (GAD67) expression in schizophrenia and bipolar disorder: a postmortem brain study. Arch Gen Psychiatry 57: 1061–1069.

83. Puvvada KC, Summerfelt A, Du X, Krishna N, Kochunov P, Rowland LM, et al. (2018): Delta Vs Gamma Auditory Steady State Synchrony in Schizophrenia. Schizophr Bull 44: 378–387.

84. Spencer KM, Niznikiewicz MA, Nestor PG, Shenton ME, McCarley RW (2009): Left auditory cortex gamma synchronization and auditory hallucination symptoms in schizophrenia. BMC Neurosci 10: 85.

85. Merritt K, McGuire PK, Egerton A, 1H-MRS in Schizophrenia Investigators, Aleman A, Block W, et al. (2021): Association of Age, Antipsychotic Medication, and Symptom Severity in Schizophrenia With Proton Magnetic Resonance Spectroscopy Brain Glutamate Level: A Mega-analysis of Individual Participant-Level Data. JAMA Psychiatry 78: 667–681.

86. Wannan CMJ, Nelson B, Addington J, Allott K, Anticevic A, Arango C, et al. (2024): Accelerating Medicines Partnership® Schizophrenia (AMP® SCZ): Rationale and study design of the largest global prospective cohort study of clinical high risk for psychosis. Schizophr Bull 50: 496–512.

87. Karvelis P, Paulus MP, Diaconescu AO (2023): Individual differences in computational psychiatry: A review of current challenges. Neurosci Biobehav Rev 148: 105137.

88. Kirkpatrick B, Messias E, Harvey PD, Fernandez-Egea E, Bowie CR (2008): Is schizophrenia a syndrome of accelerated aging? Schizophr Bull 34: 1024–1032.

89. Koutsouleris N, Davatzikos C, Borgwardt S, Gaser C, Bottlender R, Frodl T, et al. (2014): Accelerated brain aging in schizophrenia and beyond: a neuroanatomical marker of psychiatric disorders. Schizophr Bull 40: 1140–1153.

90. Kubota M, van Haren NEM, Haijma SV, Schnack HG, Cahn W, Hulshoff Pol HE, Kahn RS (2015): Association of IQ Changes and Progressive Brain Changes in Patients With Schizophrenia. JAMA Psychiatry 72: 803–812.

91. Mathalon DH, Ford JM, Rosenbloom M, Pfefferbaum A (2000): P300 reduction and prolongation with illness duration in schizophrenia. Biol Psychiatry 47: 413–427.

92. O’Donnell BF, Faux SF, McCarley RW, Kimble MO, Salisbury DF, Nestor PG, et al. (1995): Increased rate of P300 latency prolongation with age in schizophrenia. Electrophysiological evidence for a neurodegenerative process. Arch Gen Psychiatry 52: 544–549.

93. Mori Y, Kurosu S, Hiroyama Y, Niwa S-I (2007): Prolongation of P300 latency is associated with the duration of illness in male schizophrenia patients. Psychiatry Clin Neurosci 61: 471–478.

94. Legon W, Punzell S, Dowlati E, Adams SE, Stiles AB, Moran RJ (2016): Altered Prefrontal Excitation/Inhibition Balance and Prefrontal Output: Markers of Aging in Human Memory Networks. Cereb Cortex 26: 4315–4326.

95. Saberi A, Wischnewski KJ, Jung K, Lotter LD, Schaare HL, Banaschewski T, et al. (2024, June 18): Adolescent maturation of cortical excitation-inhibition balance based on individualized biophysical network modeling. BioRxiv. 10.1101/2024.06.18.599509

