## Supplement for "Excitation/inhibition imbalance and conversion to psychosis in the clinical high risk syndrome: Biophysical modeling finds reduced pyramidal cell excitability across EEG paradigms"

**Affiliations:**

**Corresponding author:**

Julia Rodriguez-Sanchez

### Supplementary Methods

#### *Participants*

A total of 269 healthy control (HC) participants were enrolled in the North American Prodrome Longitudinal Study 2 (NAPLS2). Of these, 242 provided EEG data of sufficient quality for analysis. Exclusion criteria for both clinical high risk for psychosis (CHR-P) and HC participants included any current or lifetime diagnosis of a psychotic disorder (including cluster A personality disorders for HC), an IQ below 70, or a significant central nervous system disorder (Addington et al., 2015). Participants in the HC group also had no first-degree relatives with a psychotic disorder and were not on antipsychotic medication (Addington et al., 2015).

All data were collected between May 2009 and September 2014. The study was approved by the institutional review board at each site. Adult participants provided written informed consent, and minors provided written assent, with parents providing written consent.

#### *Stimuli and task*

All auditory stimuli were presented binaurally over insert earphones (ER1-A Etymotic; Etymotic Research; https://www.etymotic.com).

The MMN paradigm consisted of 1794 trials including 1524 (85%) standard, 90 (5%) duration-deviant, 90 (5%) pitch-deviant, and 90 (5%) double-deviant tones. Trials were divided into three blocks. Standard and deviant stimuli consisted of 50 ms (standard, pitch-deviant) or 100 ms (duration-deviant, double-deviant) pure tones with a 5 ms rise and fall time. Participants completed a visual oddball task while being presented with the tones. The timing of visual stimulus presentation was jittered to prevent co-occurrence of visual oddball and MMN stimuli and to minimize correlation between MMN and visual ERP components. For more detail, please see Hamilton et al. (2022).

The P300 paradigm consisted of 450 trials including 360 (80%) standard tones, 45 (10%) target tones and 45 (10%) task-irrelevant novel sounds with a stimulus onset asynchrony of 1250 ms. Trials were divided into three blocks. Standard and target stimuli consisted of 50 ms pure tones with a 5 ms rise and fall time, while novel stimuli comprised natural and artificial sounds, such as bird calls and synthesized sounds, varying in duration (with an average of 250 ms) and in rise and fall time. Participants were asked to respond only to the target tones via a button press. For more detail, please see Hamilton et al. (2019).

#### *Data preprocessing: MMN*

As detailed in Hamilton et al. (2022), continuous EEG data were re-referenced to an average of the mastoid electrodes and high-pass filtered at 1 Hz before being segmented into 1-second epochs (-500 to 500 ms). Vertical and horizontal electro-oculogram electrodes were placed above/below the right eye and at the outer canthus of each eye to correct for blinks and saccades. Artefacts were corrected using a regression method (Gratton, Coles & Donchin, 1983) implemented in Brain Vision Analyzer (https://brainvision.com/products/analyzer-2/). Baseline correction was applied from -50 to 0 ms. Outlier electrodes within single trials were identified using Fully Automated Statistical Thresholding for EEG artifact Rejection (FASTER) (Nolan et al., 2010) criteria and interpolated using the spherical spline method implemented in EEGLAB. Bad epochs were rejected based on amplitude thresholds; specifically, epochs with amplitudes greater than ±100 µV in electrodes AF3, AF4, F3, Fz, F4, FC1, FC2, FC5, FC6, C3, Cz, or C4 were discarded. There were no group differences in the median number of good trials (Hamilton et al., 2022). Average ERPs were calculated for each condition using a sorted averaging method (Rahne, von Specht & Muhler, 2008) and low-pass filtered at 30 Hz.

Due to channel labelling and timing errors during data acquisition, channel labels for recordings from the Harvard site underwent manual editing, while event latencies for recordings from the UCSD site collected after April 27, 2011, and the Yale site collected after July 26, 2010, were adjusted by 10 ms.

#### *Data preprocessing: P300*

As detailed in Hamilton et al. (2019), continuous EEG data were re-referenced to an average of the mastoid electrodes and high-pass filtered at 0.1 Hz before being segmented into 3-second epochs (-1000 to 2000 ms). Vertical and horizontal electro-oculogram electrodes were placed above/below the right eye and at the outer canthus of each eye to correct for blinks and saccades. Artefacts were corrected using FASTER (Nolan et al., 2010), modified to include canonical correlation analysis for additional denoising of single trial data (De Clercq et al., 2006). Baseline correction was applied from -100 to 0 ms. Average ERPs were calculated for each condition and low-pass filtered at 30 Hz. Manual adjustments were made for 108 channel/timing errors at specific sites, as described above for the MMN paradigm.

#### *Software*

All analyses were conducted using the open-source software Statistical Parametric Mapping (SPM12; versions 7219 for model fitting and 7771 for group-level analyses; Wellcome Trust Centre for Neuroimaging, London, UK, https://www.fil.ion.ucl.ac.uk/spm) in MATLAB (R2018b for model fitting and R2019b for sensor-level and group-level analyses; The MathWorks Inc., Natick, MA; https://www.mathworks.com).

#### *Source localization*

Sources of the MMN include the primary auditory cortex (A1), superior temporal gyrus (STG) and inferior frontal gyrus (IFG), based on extensive work using EEG, magnetoencephalography, functional magnetic resonance imaging (fMRI) and simultaneous EEG-fMRI (Doeller et al., 2003; Garrido et al., 2009; Molholm et al., 2005; Opitz et al., 2002). Consistent with previous dynamic causal modeling (DCM) studies (e.g., Adams et al., 2022; Garrido et al., 2008), we modeled activity in these cortical areas with prior locations at [-42 -22 7] (left A1), [46 -14 8] (right A1), [-61 -32 8] (left STG), [59 -25 8] (right STG), [-46 20 8] (left IFG), [46 20 8] (right IFG), in MNI coordinates.

Sources of the P3b subcomponent have been typically localized to temporal-parietal regions, but its specific neural generators have not yet been fully characterized (Hamilton et al., 2024). To establish prior source locations for modeling, we used multiple sparse priors in SPM12 to test two different networks based on a meta-analysis of 75 oddball paradigm positron emission tomography and fMRI studies (Kim, 2014) against unconstrained source reconstruction (see also Hauke et al., 2025). Network 1 comprised bilateral auditory input sources in the STG, along with four bilateral sources of the ventral attention network: temporoparietal junction, anterior insula, anterior middle frontal gyrus, and anterior cingulate cortex. Network 2 comprised bilateral auditory input sources in the STG, alongside two bilateral sources from the dorsal attention network: inferior frontal junction (IFJ) and inferior parietal sulcus (IPS).

Source reconstructions were performed using EEG responses averaged across all HC data using the 32 channels that were common to both 32- and 64-channel systems and assessed based on their model evidence (approximated by the free energy [F]). Both network 1 (F=-643.8) and network 2 (F=-623.7) outperformed the unconstrained source reconstruction (F=-673.5), with network 2 showing the highest model evidence. This 6-region model was strongly favored in model comparison (log Bayes factor: 49.20) and explained 94.31% of the variance, compared to 95.14% for the unconstrained source reconstruction (Hauke et al., 2025). We therefore modeled activity in network 2, with prior locations at [-61 -32 8] (left STG), [59 -25 8] (right STG), [-56 7 29] (left IFJ), [50 8 30] (right IFJ), [-33 -42 64] (left IPS), [33 -42 64] (right IPS).

Importantly, although source reconstruction is not typically recommended with 32-channel EEG data, the source localization presented here was performed solely to verify that the predefined P3b network was indeed accounting for a sufficient proportion of the signal variance. Our DCM analyses did not involve estimating source locations. Instead, sources were constrained *a priori* based on the literature, which renders the inverse problem tractable.

#### *Model priors*

The default priors of the canonical microcircuit model were originally optimized for fitting high-frequency (gamma-band) activity (Pinotsis et al., 2013). It was therefore important to confirm that the synaptic time constants ($\tau$) of superficial (sp) and deep (dp) pyramidal cells, spiny stellate (ss) cells and inhibitory interneurons (ii), as well as the slope of the sigmoid output firing rate function ($S$), were appropriate for modeling the lower-frequency responses (theta and alpha range; Lee et al., 2017) examined in this study. To test this, we conducted a grid search for these parameters spanning the following grids: $\tau_{ss}$: [2 16 32 64] ms, $\tau_{ii}$: [2 16 32] ms, $\tau_{sp}$: [2 16 32 64 128] ms, $\tau_{dp}$: [2 16 32 64 128] ms and $S$ : [-2 -1 0 1 2]. The solutions were constrained to physiologically plausible parameter ranges, defined as [1 60] ms for spiny stellate cells (Markram et al., 2015; Ramaswamy et al., 2015; Stern, Edwards, & Sakmann, 1992; Sun, Huguenard, & Prince, 2006), [1 30] ms for inhibitory interneurons (Bartos et al., 2001; Hájos, & Mody, 1997) and [1 200] ms for pyramidal cells. The $\tau$ range for pyramidal cells was motivated by evidence that NMDA receptors on these neurons (which may be captured implicitly) can have time constants of around 40-200 ms (Hauke et al., 2025; Wang et al., 2008; Zhang et al., 2016). The priors identified in this grid search are reported in **Table 2** of the main manuscript**.**

To obtain suitable priors for the inversion of individual participants' data, we computed the grand mean across all participants (HC and CHR-P) and fitted the model to these grand-averaged responses. Posterior expectations of all parameters from the grand mean model with the highest model evidence across two large datasets (Oloye et al., 2025; this one) and with time constants that met the physiological constraints (Hauke et al., 2025) were used as empirical priors. These empirical priors for model inversions on single-subject data are provided for future studies of CHR-P in the code repository. For studies focused on HC or other patient groups, please see Hauke et al. (2025), who report empirical priors derived from HC only.

In the present study, the grand mean was computed from the entire NAPLS2 dataset, including all HC and CHR-P participants regardless of clinical outcome, to maximize the signal-to-noise ratio and minimize biasing the results towards finding group effects. Indeed, given that the priors were derived from data across all groups, group comparisons should not be biased by this procedure. Additionally, excluding any individual participant from the grand mean across several hundred participants is unlikely to alter the grand mean waveform. Note that, in a large dataset like this, it is not necessary to leave a participant out of the grand mean when deriving empirical priors to estimate an individual's model parameters (as is necessary in smaller datasets).

***Model inversion***

We inverted the model using the standard DCM inversion scheme of SPM that maximizes the free energy approximation to log model evidence, but replaced the default SPM integrator with a recently introduced Euler integration scheme (Schöbi et al., 2021) which is openly available as part of the Translational Algorithms for Psychiatry-Advancing Science's code toolbox (TAPAS version R09.2020; Frässle et al., 2021; https://www.translationalneuromodelling.org/tapas). This change was made to improve the robustness of the integration, which can lead to non-negligible integration errors under certain circumstances, for example for long delays (Schöbi et al., 2021). This new model termed euler_fx_cmc_ei_v1 using the Euler integrator (but also default SPM integrators) is publicly available (Hauke et al., 2025; https://github.com/daniel-hauke/dcm_ei).

***Age and sex matching procedure***

Supplementary analyses explored differences in superficial pyramidal cell and inhibitory interneuron function in CHR-Converters compared to HC. Participants in the CHR-Converter group were significantly younger than HC ($p<.05$); given known effects of age on MMN and P300 amplitude (e.g., Schiff et al., 2008), each CHR-Converter was matched to a HC to account for potential confounding effects. Matching was performed separately for the passive and active oddball tasks. For each CHR-Converter, we selected one HC of the same sex and nearest age, without replacement. After matching, there were no significant differences between groups in age or sex for either task (all $p>.05$).

Demographic characteristics of the matched CHR-Converter and HC groups, along with those of the CHR-Persistent and CHR-Remitter groups, are shown in **Tables S1** and **S2**.

#### *Group-level analysis*

Parametric empirical Bayes (PEB) is a hierarchical Bayesian framework that enables the estimation of group-level effects by leveraging prior distributions of individual subjects' parameters (Friston et al., 2016; Zeidman et al., 2019). Unlike classical statistical tests, it considers both the expectation of and covariance between parameters; put simply, parameters estimated with high confidence contribute more to the group-level inferences than those with low confidence.

In PEB, each estimated effect (PEB.Ep) is associated with a posterior probability (PEB.Pp) that the effect has diverged from its prior expectation of zero. For instance, with a posterior probability of 95% for a group effect, we are 95% confident that this first-level DCM parameter (e.g., ) is different across groups. A Bayesian framework thus bears several advantages: it allows to take uncertainty of the parameter estimates into account, it allows to estimate the confidence in the absence of an effect (null hypothesis testing) and it does not require thresholding the results. However, to provide a description of the results that is more intuitive and in line with previous (non-Bayesian) thresholds, we consider effects to be significant if the posterior probability exceeds 95% (analogous to p-values smaller than .05) (Kass & Raftery, 1995).

### Supplementary Results

In the passive oddball paradigm, we found no group differences in superficial pyramidal cell (P=.92) or inhibitory interneuron (P=.56) excitability between CHR-Converters and HC (**Figure S5A**). In contrast, in the active oddball paradigm, there was increased pyramidal cell (P>.99) and inhibitory interneuron self-inhibition (i.e., decreased excitability; P>.95) in CHR-Converters compared to HC (**Figure S5B**).

Previous analyses of these data examined clinical outcomes across a broader set of subgroups, including CHR-P individuals who showed symptom persistence or symptom and/or syndrome progression at the 24-month assessment, defined as "continuing to meet Criteria of Psychosis-Risk Syndromes or experiencing attenuated positive symptoms over the previous 4 weeks" (CHR-Persistent) (Hamilton et al., 2019, 2022). Notably, participants in the CHR-Persistent group were found to differ significantly from HC and CHR-Remitters in P300 amplitude, with no differences relative to CHR-Converters (Hamilton et al., 2019). To facilitate comparison with previous work, we conducted additional analyses comparing CHR-Converters versus CHR-Nonconverters (including CHR-Remitters plus CHR-Persistent). We also modeled the CHR-Converters plus CHR-Persistent versus CHR-Remitters effect on P300.

In the passive oddball paradigm, CHR-Converters showed increased pyramidal cell (P>.95) and inhibitory interneuron (P>.99) self-inhibition compared to CHR-Nonconverters (**Figure S5C**). In contrast, no differences were observed between CHR-Converters and CHR-Nonconverters in the active oddball paradigm (both P=.85; **Figure S5D**). Instead, increased pyramidal cell and inhibitory interneuron self-inhibition (both P>.95) were observed in CHR-Converter plus CHR-Persistent participants compared to CHR-Remitters (**Figure S5E**).

Given the presence of inhibitory interneuron hypoexcitability alongside pyramidal cell hypoexcitability in these contrasts, one might question whether there is equal evidence for each being a primary pathology in CHR-P individuals. The simulations indicate that interneuron hypoexcitability ought to increase MMN and P300 amplitudes (**Figure 3**, main paper), however – opposite to the group average effects in CHR individuals – so it seems most likely that this interneuron dysfunction is a compensatory effect, and/or a property of a smaller subgroup within the CHR-P. Analysis of psychosis 'biotypes' (Clementz et al., 2016) may inform this question further.

### Supplementary Tables

**Table S1. Demographic and clinical characteristics for the passive oddball task.**

| Demographic | Healthy Control (n=77) | CHR-Converter (n=77) | CHR-Persistent (n=144) | CHR-Remitter (n=94) | Statistical analysis | Post hoc contrast |
|---|---|---|---|---|---|---|
| **Age [mean (SD)]** | 19 (3.6) | 19 (3.6) | 20 (4.6) | 19 (4.4) | **F=3.08, p=.028** | - |
| **Female [n (%)]** | 29 (37.7) | 29 (37.7) | 60 (41.7) | 47 (50.0) | χ2=3.66, p=.300 | - |
| **CHR-P syndrome [n (%)]** [a] | | | | | | |
| APSS | - | 73 (94.8) | 136 (94.4) | 88 (93.6) | χ2=0.12, p=.940 | - |
| BIPS | - | 8 (10.4) | 2 (1.4) | 1 (1.1) | **χ2=14.41, p<.001** | **CHR-Converter v CHR-Persistent: p=.002**<br>**CHR-Converter v CHR-Remitter: p=.007** |
| GRDS | - | 14 (18.2) | 16 (11.1) | 11 (11.7) | χ2=2.42, p=.298 | - |
| **Current antipsychotic use [n (%)]** | - | 23 (29.9) | 32 (22.2) | 16 (17.0) | χ2=4.02, p=.134 | - |
| **SOPS [mean (SD)]** | | | | | | |
| Positive | - | 13.4 (3.8) | 12.4 (3.8) | 10.7 (4.4) | **F=10.50, p<.001** | **CHR-Converter v CHR-Remitter: p<.001**<br>**CHR-Persistent v CHR-Remitter: p=.003** |
| Negative | - | 12.3 (6.3) | 11.2 (6.4) | 10.9 (6.0) | F=1.10, p=.333 | - |
| Disorganization | - | 6.1 (3.9) | 4.9 (3.1) | 4.1 (2.9) | **F=7.54, p<.001** | **CHR-Converter v CHR-Persistent: p=.031**<br>**CHR-Converter v CHR-Remitter: p<.001** |
| General | - | 9.5 (4.3) | 8.1 (4.3) | 7.8 (4.4) | **F=3.82, p=.023** | **CHR-Converter v CHR-Remitter: p=.025** |

One-way analysis of variance (ANOVA) was used to assess group differences in age and Scale of Psychosis-Risk Symptoms (SOPS; McGlashan, Walsh, & Woods, 2010) ratings; χ2 tests were used to assess group differences in sex, clinical high risk for psychosis (CHR-P), medication and available data between CHR-P participants who subsequently converted to psychosis (CHR-Converter), had persistent symptoms (CHR-Persistent), or whose symptoms remitted (CHR-Remitter) and a subset of healthy controls age and sex matched to the CHR-Converter group. Values are reported as mean (standard deviation [SD]) for continuous variables and as count [n] (percentage [%]) for categorical variables. [a]Criteria for attenuated psychotic symptom syndrome (APSS), brief intermittent psychotic syndrome (BIPS), and genetic risk and deterioration syndrome (GRDS) are not mutually exclusive.

**Table S2. Demographic and clinical characteristics for the active oddball task.**

| Demographic | Healthy Control (n=73) | CHR-Converter (n=73) | CHR-Persistent (n=135) | CHR-Remitter (n=89) | Statistical analysis | Post hoc contrast |
|---|---|---|---|---|---|---|
| **Age [mean (SD)]** | 19 (3.6) | 18 (3.6) | 20 (4.6) | 19 (4.5) | **F=2.71, p=.045** | - |
| **Female [n (%)]** | 27 (37.0) | 27 (37.0) | 58 (43.0) | 45 (50.6) | χ2=3.57, p=.312 | - |
| **CHR-P syndrome [n (%)]** [a] | | | | | | |
| APSS | - | 70 (95.9) | 127 (94.1) | 83 (93.3) | χ2=0.53, p=.766 | - |
| BIPS | - | 8 (11.0) | 2 (1.5) | 1 (1.1) | **χ2=14.30, p<.001** | **CHR-Converter v CHR-Persistent: p=.002**<br>**CHR-Converter v CHR-Remitter: p=.007** |
| GRDS | - | 12 (16.4) | 15 (11.1) | 11 (12.4) | χ2=1.23, p=.542 | - |
| **Current antipsychotic use [n (%)]** | - | 22 (30.1) | 31 (23.0) | 16 (18.0) | χ2=3.33, p=.189 | - |
| **SOPS [mean (SD)]** | | | | | | |
| Positive | - | 13.2 (4.0) | 11.8 (4.1) | 9.7 (4.6) | **F=14.2, p<.001** | **CHR-Converter v CHR-Persistent: p=.044**<br>**CHR-Converter v CHR-Remitter: p<.001**<br>**CHR-Persistent v CHR-Remitter: p=.001** |
| Negative | - | 12.3 (6.3) | 11.4 (6.5) | 11.1 (5.9) | F=0.9, p=.423 | - |
| Disorganization | - | 6.1 (4.0) | 5.0 (3.1) | 4.2 (3.0) | **F=6.8, p=.001** | **CHR-Converter v CHR-Remitter: p<.001** |
| General | - | 9.5 (4.3) | 8.1 (4.3) | 8.0 (4.5) | **F=3.32, p=.037** | - |

One-way analysis of variance (ANOVA) was used to assess group differences in age and Scale of Psychosis-Risk Symptoms (SOPS; McGlashan, Walsh, & Woods, 2010) ratings; χ2 tests were used to assess group differences in sex, clinical high risk for psychosis (CHR-P), medication and available data between CHR-P participants who subsequently converted to psychosis (CHR-Converter), had persistent symptoms (CHR-Persistent), or whose symptoms remitted (CHR-Remitter) and a subset of healthy controls age and sex matched to the CHR-Converter group. Values are reported as mean (standard deviation [SD]) for continuous variables and as count [n] (percentage [%]) for categorical variables. [a]Criteria for attenuated psychotic symptom syndrome (APSS), brief intermittent psychotic syndrome (BIPS), and genetic risk and deterioration syndrome (GRDS) are not mutually exclusive.

### Supplementary Figures

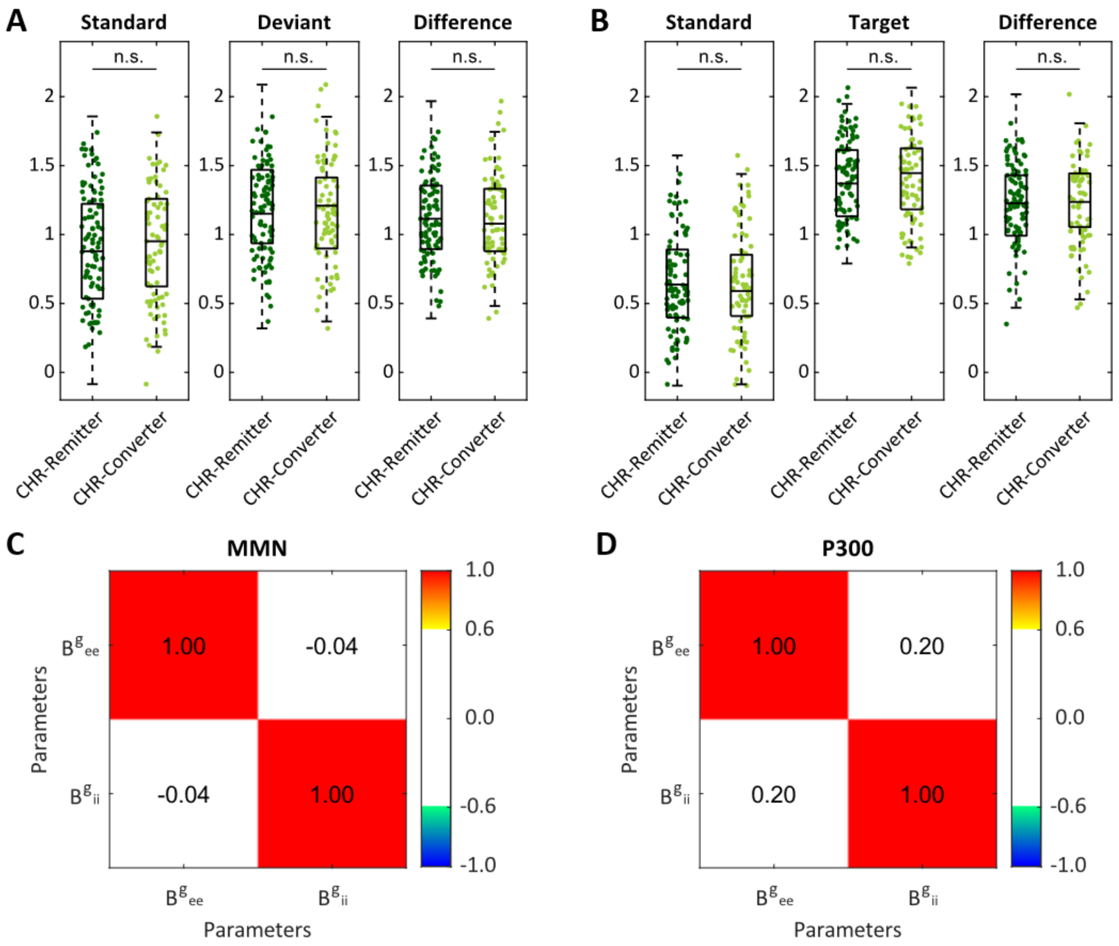


**Figure S1. Model fit and diagnostics.** Panels **(A)** and **(B)** show the distribution of Fisher *z*-transformed correlation coefficients across clinical high risk for psychosis (CHR-P) participants who subsequently converted to psychosis (CHR-Converters) and those whose symptoms remitted (CHR-Remitters) for **(A)** the MMN paradigm and **(B)** the P300 paradigm. Each dot represents an individual participant. Group differences were tested using Mann-Whitney U tests (n.s., not significant). Panels **(C)** and **(D)** show correlation matrices with between-subject parameter correlations to illustrate parameter identifiability for **(C)** the MMN paradigm and **(D)** the P300 paradigm.

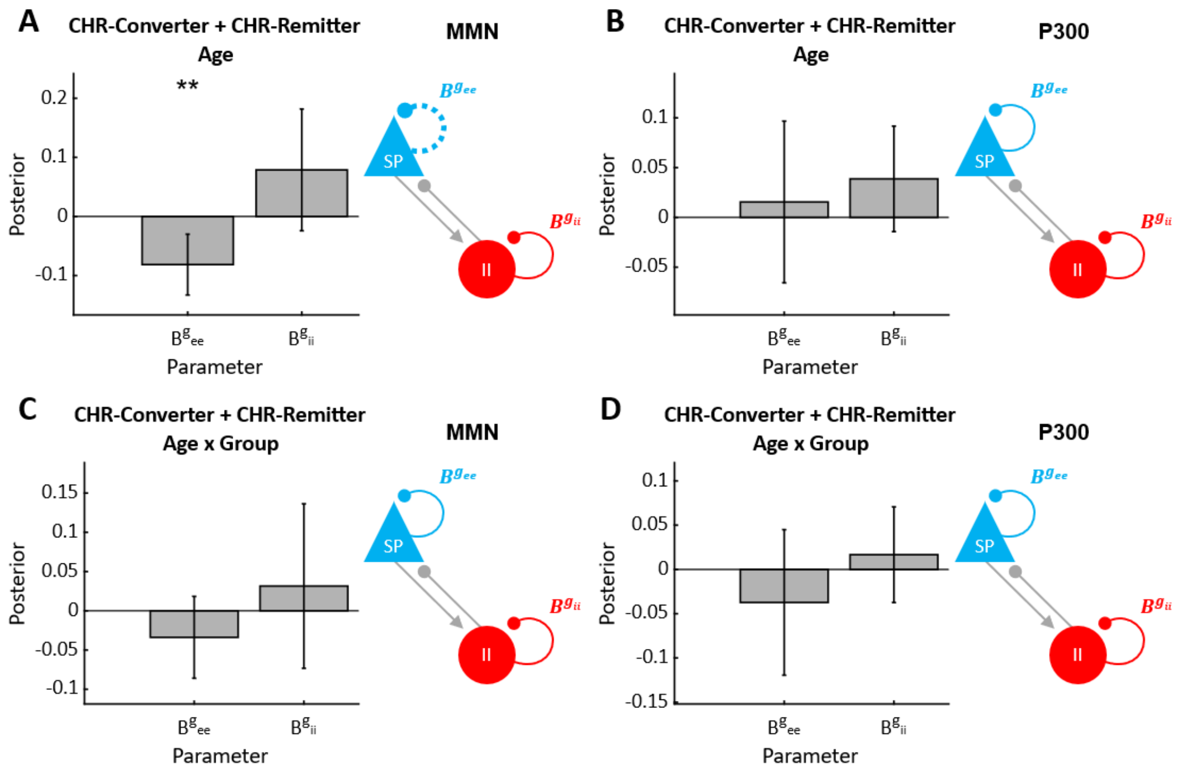


**Figure S2. Age effects on mismatch negativity (MMN) and P300 model parameters.** Panels **(A)** and **(B)** show the main effect of age on condition-specific modulation of superficial pyramidal cell self-inhibition ($B^{g_{ee}}$) and inhibitory interneuron self-inhibition ($B^{g_{ii}}$) in CHR-Converters and CHR-Remitters for **(A)** the MMN paradigm and **(B)** the P300 paradigm. Panels **(C)** and **(D)** show the age-by-group interaction effect, estimated from a model that also included the main effects of age and group. Across all panels, error bars represent the 95% Bayesian confidence intervals. Asterisks indicate a significant effect with a posterior probability of P>.95 (*) or P>.99 (**). In the microcircuit diagrams, parameter decreases are illustrated with dotted lines.

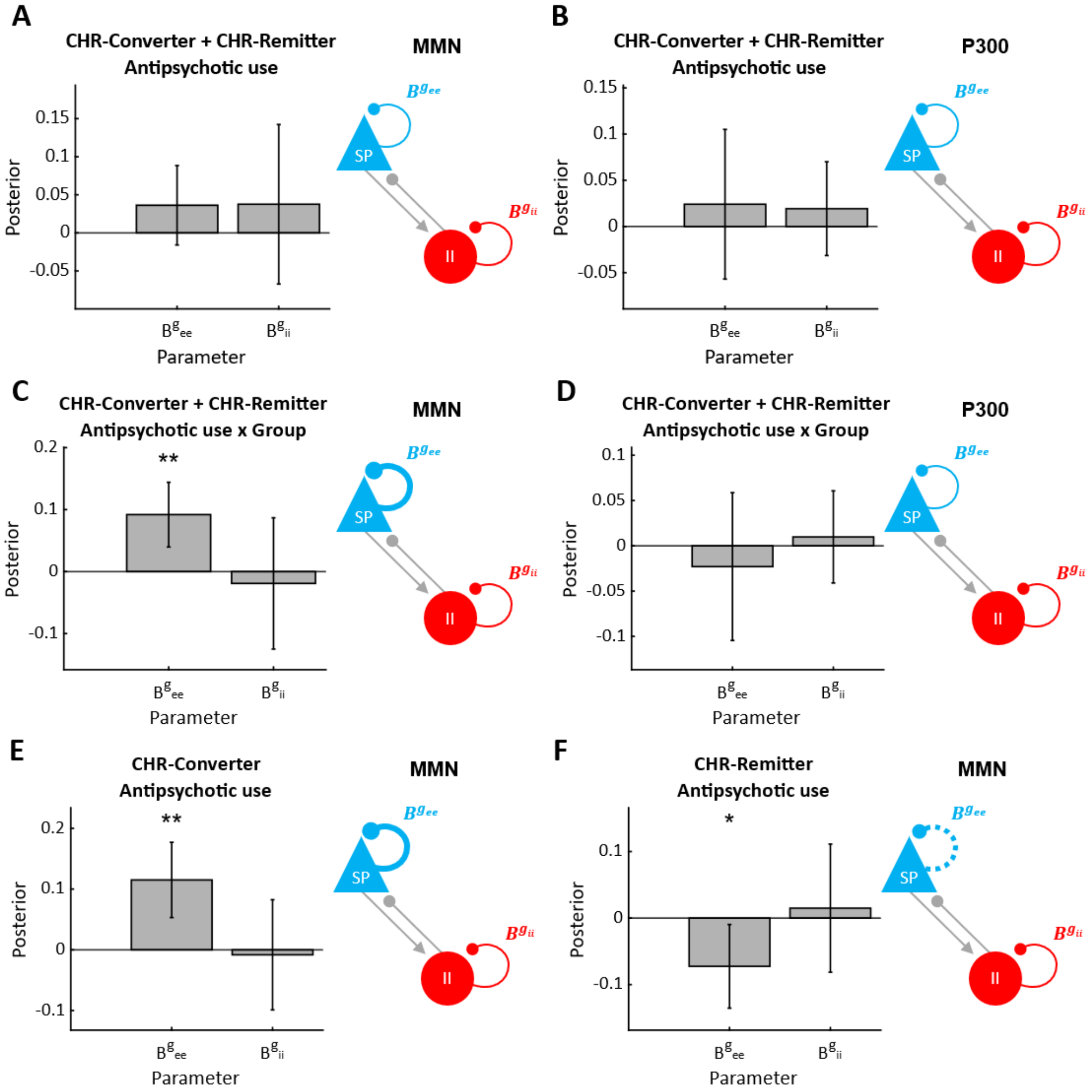


**Figure S3. Antipsychotic use effects on mismatch negativity (MMN) and P300 model parameters.** Panels **(A)** and **(B)** show the effects of antipsychotic use on condition-specific modulation of superficial pyramidal cell self-inhibition ($B^{g_{ee}}$) and inhibitory interneuron self-inhibition ($B^{g_{ii}}$) in clinical high risk for psychosis (CHR-P) participants who subsequently converted to psychosis (CHR-Converters) and those whose symptoms remitted (CHR-Remitters) for **(A)** the MMN paradigm and **(B)** the P300 paradigm. Panels **(C)** and **(D)** show the antipsychotic use-by-group interaction effect. Panels **(E)** and **(F)** show the effect of antipsychotic use in **(E)** CHR-Converters and **(F)** CHR-Remitters for the MMN paradigm. Across all panels, error bars represent the 95% Bayesian confidence intervals. Asterisks indicate a significant effect with a posterior probability of (*) P>.95 or (**) P>.99. In the microcircuit diagrams, parameter increases are illustrated with thick lines, and parameter decreases with dotted lines.

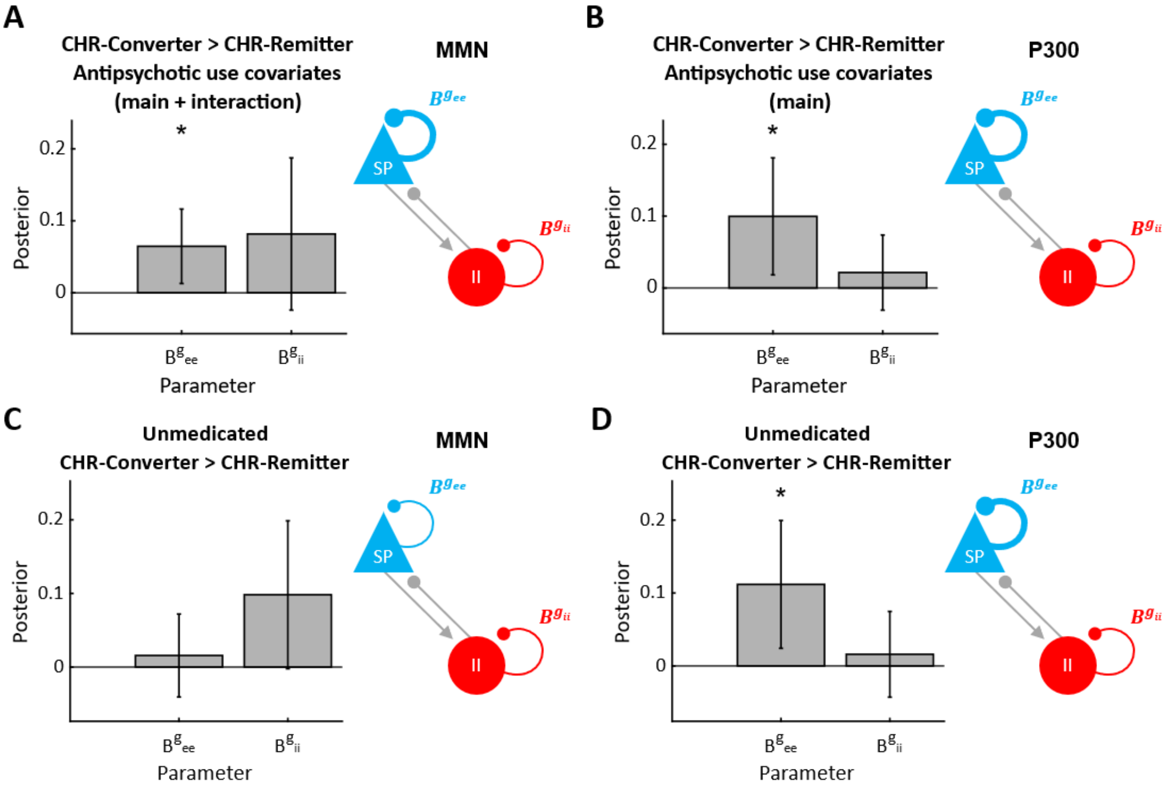


**Figure S4. Group-level analysis of mismatch negativity (MMN) and P300 model parameters: antipsychotic use covariates and unmedicated subsample.** Panels **(A)** and **(B)** show group differences in condition-specific modulation of superficial pyramidal cell self-inhibition ($B^{g_{ee}}$) and inhibitory self-inhibition ($B^{g_{ii}}$) in clinical high risk for psychosis (CHR-P) participants who subsequently converted to psychosis (CHR-Converters) compared to those whose symptoms remitted (CHR-Remitters), with inclusion of antipsychotic use as a covariate, for **(A)** the MMN paradigm and **(B)** the P300 paradigm. In (A) the second-level model also included an antipsychotic use-by-group interaction term. Panels **(C)** and **(D)** show the same group contrasts in a subsample of unmedicated CHR-Converters and unmedicated CHR-Remitters. Across all panels, error bars represent the 95% Bayesian confidence intervals. Asterisks (*) indicate a significant group effect with a posterior probability of P>.95. In the microcircuit diagrams, parameter increases are illustrated with thick lines.

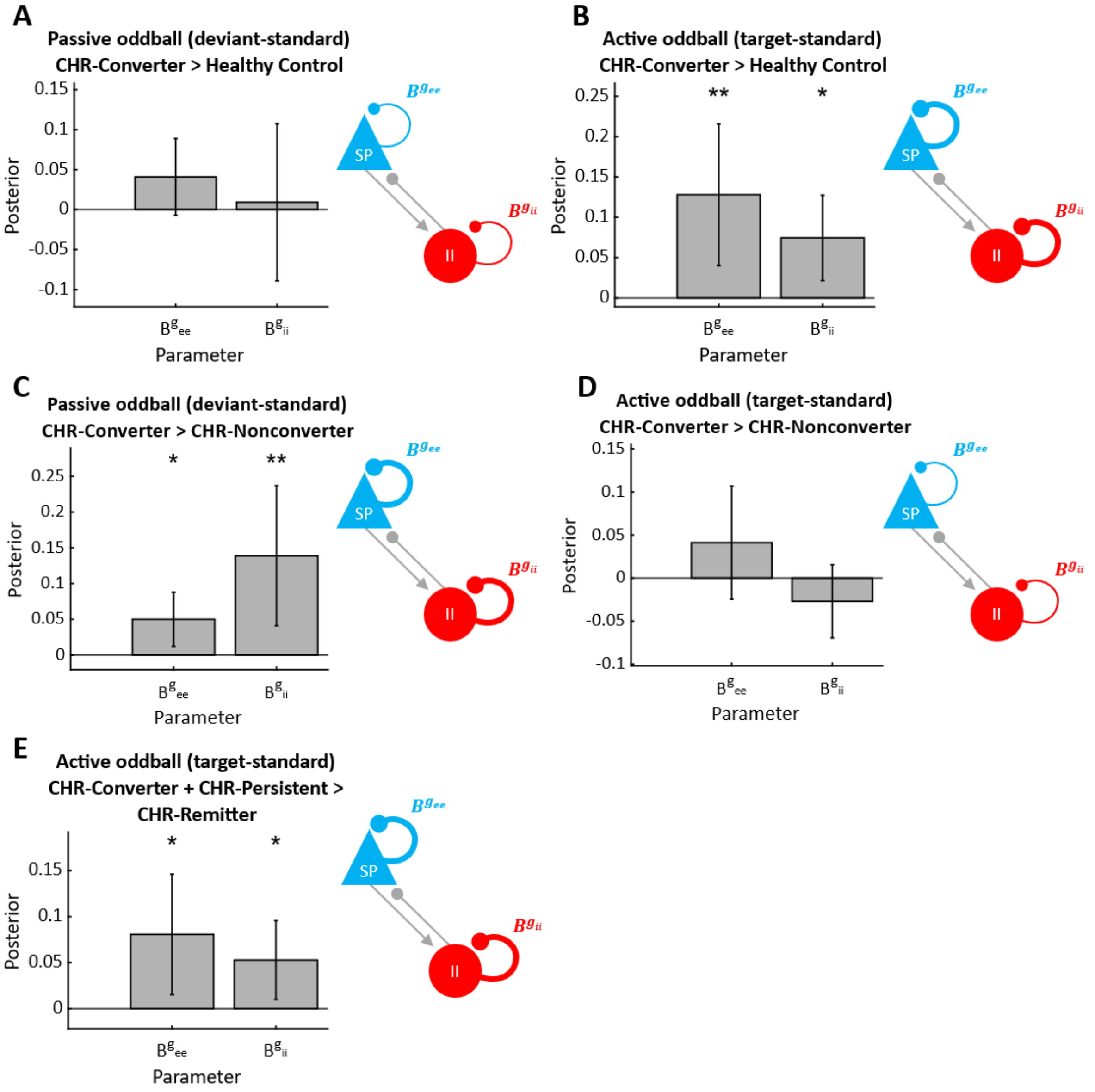


**Figure S5. Additional group-level results.** Panels **(A)** and **(B)** show group differences in condition-specific modulation of superficial pyramidal cell self-inhibition ($B^{gee}$) and inhibitory self-inhibition ($B^{gii}$) in clinical high risk for psychosis (CHR-P) participants who subsequently converted to psychosis (CHR-Converter) compared to healthy controls for **(A)** the MMN paradigm and **(B)** the P300 paradigm. **(C)** and **(D)** show the same in the CHR-Converter group compared to CHR-P participants who did not convert (CHR-Nonconverter). Panel **(E)** shows group differences in $B^{gee}$ and $B^{gii}$ for the P300 paradigm in CHR-Converter and those with persistent CHR-P symptoms (CHR-Persistent) compared to CHR-P participants who remitted (CHR-Remitters). Across all panels, error bars represent the 95% Bayesian confidence intervals. Asterisks (*) indicate a significant group effect. In the microcircuit diagrams, parameter increases are illustrated with thick lines.

**References**


Bartos, M., Vida, I., Frotscher, M., Geiger, J. R., & Jonas, P. (2001). Rapid signaling at inhibitory synapses in a dentate gyrus interneuron network. *Journal of Neuroscience*, 21(8), 2687-2698. https://doi.org/10.1523/JNEUROSCI.21-08-02687.2001

Clementz, B. A., Sweeney, J. A., Hamm, J. P., Ivleva, E. I., Ethridge, L. E., Pearlson, G. D., ... & Tamminga, C. A. (2016). Identification of distinct psychosis biotypes using brain-based biomarkers. *American Journal of Psychiatry*, 173(4), 373-384. https://doi.org/10.1176/appi.ajp.2015.14091200

De Clercq, W., Vergult, A., Vanrumste, B., Van Paesschen, W., & Van Huffel, S. (2006). Canonical correlation analysis applied to remove muscle artifacts from the electroencephalogram. *IEEE Transactions on Biomedical Engineering*, 53(12), 2583-2587. https://doi.org/10.1109/TBME.2006.879459

Gratton, G., Coles, M. G., & Donchin, E. (1983). A new method for off-line removal of ocular artifact. *Electroencephalography and Clinical Neurophysiology*, 55(4), 468-484. https://doi.org/10.1016/0013-4694(83)90135-9

Hájos, N., & Mody, I. (1997). Synaptic communication among hippocampal interneurons: properties of spontaneous IPSCs in morphologically identified cells. *Journal of Neuroscience*, 17(21), 8427-8442. https://doi.org/10.1523/JNEUROSCI.17-21-08427.1997

Hamilton, H. K., Roach, B. J., Bachman, P. M., Belger, A., Carrion, R. E., Duncan, E., Johannesen, J. K., Light, G. A., Niznikiewicz, M. A., Addington, J., Bearden, C. E., Cadenhead, K. S., Cornblatt, B. A., McGlashan, T. H., Perkins, D. O., Seidman, L. J., Tsuang, M. T., Walker, E. F., Woods, S. W., … Mathalon, D. H. (2019). Association Between P300 Responses to Auditory Oddball Stimuli and Clinical Outcomes in the Psychosis Risk Syndrome. *JAMA Psychiatry*, *76*(11), 1187–1197. https://doi.org/10.1001/jamapsychiatry.2019.2135

Hauke, D. J., Rodriguez-Sanchez, J., Oloye, H., Berndt, L., Pinotsis, D., Friston, K. J., Mathalon, D. H., Adams, R. A. (2025). A canonical microcircuit for estimating excitation/inhibition (E/I) balance. *BioRxiv.* https://doi.org/10.1101/2025.07.10.664116

Kim, H. (2014). Involvement of the dorsal and ventral attention networks in oddball stimulus processing: A meta-analysis. *Human Brain Mapping*, 35(5), 2265-2284. https://doi.org/10.1002/hbm.22326

Markram, H., Muller, E., Ramaswamy, S., Reimann, M. W., Abdellah, M., Sanchez, C. A., ... & Schürmann, F. (2015). Reconstruction and simulation of neocortical microcircuitry. Cell, 163(2), 456-492. http://dx.doi.org/10.1016/j.cell.2015.09.029

Nolan, H., Whelan, R., & Reilly, R. B. (2010). FASTER: fully automated statistical thresholding for EEG artifact rejection. *Journal of Neuroscience Methods*, 192(1), 152-162. https://doi.org/10.1016/j.jneumeth.2010.07.015

Oloye, H., Hauke, D. J., Rodriguez-Sanchez, J., Parker, D., Pearlson, G., Keshavan, M., Gershon, E., Keedy, S., Clementz, B., Tamminga, C., Adams, R. A. (2025). Computational modelling of paired clicked and auditory oddball ERP responses to probe inhibitory and excitatory cell function in schizophrenia and across transdiagnostic biotypes: Insights from the B-SNIP study. *In preparation.*

Opitz, B., Rinne, T., Mecklinger, A., von Cramon, D. Y., Schröger, E. (2002). Differential contribution of frontal and temporal cortices to auditory change detection: fMRI and ERP results. *Neuroimage,* 15, 167–174. https://doi.org/10.1006/nimg.2001.0970

Pinotsis, D. A., Schwarzkopf, D. S., Litvak, V., Rees, G., Barnes, G., & Friston, K. J. (2013). Dynamic causal modelling of lateral interactions in the visual cortex. *NeuroImage*, 66, 563–576. https://doi.org/10.1016/j.neuroimage.2012.10.078

Rahne, T., von Specht, H., & Mühler, R. (2008). Sorted averaging—application to auditory event-related responses. *Journal of Neuroscience Methods*, 172(1), 74-78. https://doi.org/10.1016/j.jneumeth.2008.04.006

Ramaswamy, S., Courcol, J. D., Abdellah, M., Adaszewski, S. R., Antille, N., Arsever, S., ... & Markram, H. (2015). The neocortical microcircuit collaboration portal: a resource for rat somatosensory cortex. *Frontiers in Neural Circuits*, 9, 44. https://doi.org/10.3389/fncir.2015.00044

Schiff, S., Valenti, P., Andrea, P., Lot, M., Bisiacchi, P., Gatta, A., & Amodio, P. (2008). The effect of aging on auditory components of event-related brain potentials. *Clinical Neurophysiology*, 119(8), 1795-1802. https://doi.org/10.1016/j.clinph.2008.04.007

Sun, Q. Q., Huguenard, J. R., & Prince, D. A. (2006). Barrel cortex microcircuits: thalamocortical feedforward inhibition in spiny stellate cells is mediated by a small number of fast-spiking interneurons. *Journal of Neuroscience*, 26(4), 1219-1230. https://doi.org/10.1523/JNEUROSCI.4727-04.2006

Stern, P., Edwards, F. A., & Sakmann, B. (1992). Fast and slow components of unitary EPSCs on stellate cells elicited by focal stimulation in slices of rat visual cortex. *The Journal of Physiology*, 449(1), 247-278. https://doi.org/10.1113/jphysiol.1992.sp019085

Wang, H., Stradtman III, G. G., Wang, X. J., & Gao, W. J. (2008). A specialized NMDA receptor function in layer 5 recurrent microcircuitry of the adult rat prefrontal cortex. *Proceedings of the National Academy of Sciences*, 105(43), 16791-16796. https://doi.org/10.1073/pnas.0804318105

Zhang, W. B., Ross, P. J., Tu, Y., Wang, Y., Beggs, S., Sengar, A. S., ... & Salter, M. W. (2016). Fyn Kinase regulates GluN2B subunit-dominant NMDA receptors in human induced pluripotent stem cell-derived neurons. *Scientific Reports*, 6(1), 23837. https://doi.org/10.1038/srep23837
